# The impact of female sex on anaesthetic awareness, depth and emergence: A systematic review and meta-analysis

**DOI:** 10.1101/2023.03.14.23287147

**Authors:** Hannah E. Braithwaite, Thomas Payne, Nicholas Duce, Jessica Lim, Tim McCulloch, John Loadsman, Kate Leslie, Angela C Webster, Robert D. Sanders

**Affiliations:** Department of Anaesthetics, Royal Prince Alfred Hospital, Sydney Local Health District, NSW, Australia; Central Clinical School, Faculty of Medicine and Health, The University of Sydney, NSW, Australia; Institute of Academic Surgery, Royal Prince Alfred Hospital, Sydney Local Health District, NSW, Australia; Department of Critical Care, Melbourne Medical School, University of Melbourne, Melbourne, VIC, Australia; Central Clinical School, Faculty of Medicine, Dentistry and Health Sciences, Monash University, Melbourne, VIC, Australia; Department of Anaesthesia and Pain Management, Royal Melbourne Hospital, Parkville, VIC, Australia; Sydney School of Public Health, Faculty of Medicine and Health, University of Sydney, Sydney, NSW, Australia; Centre for Transplant and Renal Research, Westmead Hospital, Sydney, NSW, Australia; National Health and Medical Research Council Clinical Trials Centre, Faculty of Medicine and Health, The University of Sydney, NSW, Australia

**Keywords:** sex, general anaesthesia, intra-operative awareness, connected consciousness

## Abstract

**Background:** Accumulating evidence supports sex differences in pharmacodynamic and pharmacokinetic drug profiles. However, recommended anaesthetic drug doses are not sex-adjusted, likely due to limited studies comparing sexes. Our objective was to systematically synthesise studies of anaesthetic key performance indicators (anaesthesia awareness), and markers of relatively lighter anaesthesia, time to emergence and dosing to achieve adequate depth of anaesthesia, for females and males.

**Methods:** MEDLINE, Embase, and the Cochrane library databases. Studies were identified from inception of database to August 2^nd^, 2022. Controlled clinical trials (randomised and non-randomised) and prospective cohort studies that reported outcomes by sex for awareness with post-operative recall, connected consciousness during anaesthesia, depth of anaesthesia, and emergence from anaesthesia. Two authors undertook search, review, selection, and data abstraction. Risk of bias was assessed using the Newcastle Ottawa Scale. Results were synthesized by random effects meta-analysis where possible, or narrative form. Results were expressed as odds ratios (ORs) and mean differences (MDs) with corresponding 95% confidence intervals (CIs).

**Results:** Of the 19,749 studies identified from literature search, 66 citations of 64 studies (98,243 participants; 53,143 females and 45,100 males) were eligible for inclusion, of which 44 contributed to meta-analysis. Females had a higher incidence of awareness with post-operative recall (33 studies, OR 1.37, 95%CI 1.09 to 1.75) and connected consciousness during anaesthesia (3 studies, OR 2.09, 95% CI 1.04 to 4.23) than males. Time to emergence was faster in females than in males, including time to eye-opening (10 studies, MD -2.28 min, 95% CI -3.58 to -0.98), and time to response to command (6 studies, MD - 2.84 min, 95% CI -4.07 to -1.62). Data on depth of anaesthesia were heterogenous limiting synthesis to a qualitative review which did not identify differences by sex.

**Conclusion:** Female sex was associated with a greater incidence of anaesthetic awareness, as well as faster emergence from anaesthesia. These data suggest reappraisal of anaesthetic care, including whether similar drug dosing for females and males represents best care. Equitable outcomes for females undergoing general anaesthesia warrants strategic focus in future research.

**Systematic review registration:** PROSPERO CRD42022336087.

## INTRODUCTION

Anaesthesia is provided to hundreds of millions of people each year^1^ with remarkable safety.^2^ However, patients remain concerned about experiencing connected consciousness (involving the experience of external environmental and surgical stimuli) whilst under general anaesthesia,^3^ which can be considered a key performance indicator (KPI) of the effectiveness of anaesthesia provision. Postoperative recall of events under general anaesthesia, occurs in approximately 1-2 in 1000 anaesthetics,^4-6^ with reported rates of post-traumatic stress disorder in those affected as high as 43%.^7^ Hence, regular appraisal of anaesthesia awareness, as an essential performance indicator, is critical to advancing the field of anaesthesiology.

Steep dose-response curves for aanesthetics mean that small differences in dosing may result in ineffective anaesthesia, leading us to consider important factors that may influence anaesthetic outcomes and dosing. Sex, typically categorized in this setting as a biological binary form of female and male,^8^ is widely assumed to not affect anaesthetic drug-dosing.^9^ Product information for commonly used anaesthetic agents lacks differentiated recommendations for therapeutic dosing between females and males.^10-12^ However, recent data suggest that sex hormones alter anaesthetic requirements,^13-16^ implying potentially important pharmacodynamic or kinetic differences between females and males. Our recent multi-centre cohort study of connected consciousness during intended anaesthesia (defined by response to command during intended general anaesthesia) suggested that females may be three times more likely than males to respond to command following intubation during general anaesthesia.^17^ Interestingly, females and males received similar doses of anaesthetic drugs, implying there is a lack of consideration that female sex is a risk factor for awareness.^18^ This view is supported by a recent authoritative review of anaesthetic awareness that did not mention sex as a risk factor.^6^ While case series (including those with estimated control populations) suggest females may be more likely to have awareness with post-operative recall than males,^19^ key prospective cohort studies and trials have not indicated the same.^4,20^ However, due to the relatively low incidence of recall following anaesthesia real differences may not be evident in individual studies due to lack of power. This has perpetuated the belief that anaesthetic requirements are similar between females and males.

We conducted a systematic review of the impact of sex on anaesthesia awareness, a KPI of the effectiveness of anaesthesia provision. We included two separate definitions of anaesthesia awareness, expecting convergent findings across these endpoints. These included (1) connected consciousness as described by intraoperative responsiveness to command under intended general anaesthesia, and (2) postoperative recall of events following intended general anaesthesia. We supplemented these analyses with exploration of studies of time to emergence, as relatively lighter anaesthesia leads to faster emergence. Finally, we appraised the literature for further evidence that sex may influence doses of anaesthetics to achieve adequate depth of anaesthesia.

## METHODS

The review protocol for this systematic review with meta-analyses was registered on PROSPERO: CRD42022336087.

### Search strategy and selection criteria

Databases including MEDLINE, Embase and the Cochrane Library were searched from inception to August 2^nd^, 2022. Reference lists of included studies were reviewed to identify additional relevant studies for inclusion. Titles and abstracts were screened by one reviewer (HEB), whilst full text articles were independently screened by two reviewers (HEB, ND).

#### Discrepancies were resolved by consensus

Studies were eligible for inclusion if they were controlled trials or cohort studies performed in adults undergoing general anaesthesia. Included studies were prospective, and reported at least one of the following outcomes stratified by sex: (1) awareness with post-operative recall as assessed by the Modified Brice^21^, or similarly structured questionnaire; (2) connected consciousness during anaesthesia (defined as response to command using the isolated forearm technique [IFT]); (3) emergence from anaesthesia (including time from cessation of anaesthetic delivery to eye opening, response to command, or extubation, or duration of post-anaesthesia care unit [PACU] stay); and (4) depth of anaesthesia (as measured by processed electroencephalographic [pEEG] monitoring, clinical signs including haemodynamic changes, and Observer’s Assessment of Alertness/Sedation [OAA/S] scores). For outcomes 3 and 4, studies of sedation were also included as depth of anaesthesia and emergence parameters can be assessed across a range of drug doses.

### Data extraction and quality assessment

Bias appraisal was performed using the Newcastle Ottawa Scale cohort study tool, with scores of 6-7, 5, and ≤4 considered to represent high, fair or poor quality, respectively. Risk of bias assessments were performed independently by two reviewers (HEB, ND). Publication integrity was assessed against the REAPPRAISED checklist.^22^ Where studies had concerns identified, we performed sensitivity analyses.

Data were extracted independently by two reviewers (HEB, TP). WebPlotDigitizer^23^ was used to extract numeric data from figure format. Study authors contacted if further data were required. For the purposes of this study, sex was considered as a biological binary form of females and males. Classification of post-operative awareness with recall into confirmed or possible categories was in accordance with individual study definitions.

### Data synthesis and statistical analysis

Where quantitative analysis was not feasible (<3 studies reporting congruent outcome measures), results were reported in narrative synthesis. Meta-analyses were performed on the incidence of awareness with post-operative recall following general anaesthesia, connected consciousness, and time to emergence data. All statistical analyses were performed in R (R Studio 2022.02.1 build 461, base R 4.1.3) using the Metafor package.^24^ For dichotomous outcomes, we reported odds ratios (ORs), while for continuous outcomes we reported mean differences (MDs), both with 95% confidence intervals (CIs). Where continuous outcomes were reported in ranges or CIs, standard deviations were calculated using methods provided in the Cochrane handbook.^25^ Heterogeneity was quantified using Cochran’s Q, τ^2^, and I^2^ statistics. We considered an I^2^ of 25%, 50%, and 75% to represent low, moderate, and high heterogeneity, respectively.^26^ The restricted maximum likelihood approach was used to estimate τ^2^ in random effects meta-analyses.^27^ The Hartung–Knapp– Sidik–Jonkman adjustment to the standard errors was used for two-level random effects meta-analyses.^28-30^ Where the I^2^ > 0%, 95% prediction intervals were calculated based on critical values from a t-distribution.^31^

A fixed-effect meta-analysis, using the Peto method,^32^ was used to analyse rare outcomes, as recommended by Cochrane, including pooled data of awareness with post-operative recall.^25^ For studies of IFT responsiveness, we used a two-level random effects model. Meta-analysis of time to eye opening data was performed using a three-level random effects model, given the considerable number of dependent effect sizes. Confidence intervals were based on a *t*-distribution. Potential sources of heterogeneity were explored using meta-regression, including age, duration of surgery and type of maintenance anaesthesia, after assessment for collinearity. We selected the model that provided the best fit based on a multimodel inference method (without interactions) in the dmetar package.^33^ The model was assessed using permutation tests, robust variance estimation and traditional fit statistics. All other time to emergence meta-analyses were performed using a two-level random effects model. Several sensitivity analyses were performed, including alternative methods of meta-analysis, other estimators for τ^2^, and sequential exclusion analysis, which are included within the supplementary material.

## RESULTS

We included 66 citations of 64 studies, involving 98,243 participants (53,143 females, 45,100 males; fig 1), of which 44 studies contributed to meta-analysis. The methodology for the search strategy is included in supplementary Table 2. Three citations^34-36^ were derived from the same trial and were thus nested under the initial study publication.^34^ Four hundred and seven studies (78%) reviewed in full text were excluded due to lack of sex stratification of reported outcomes.

**Figure 1.**
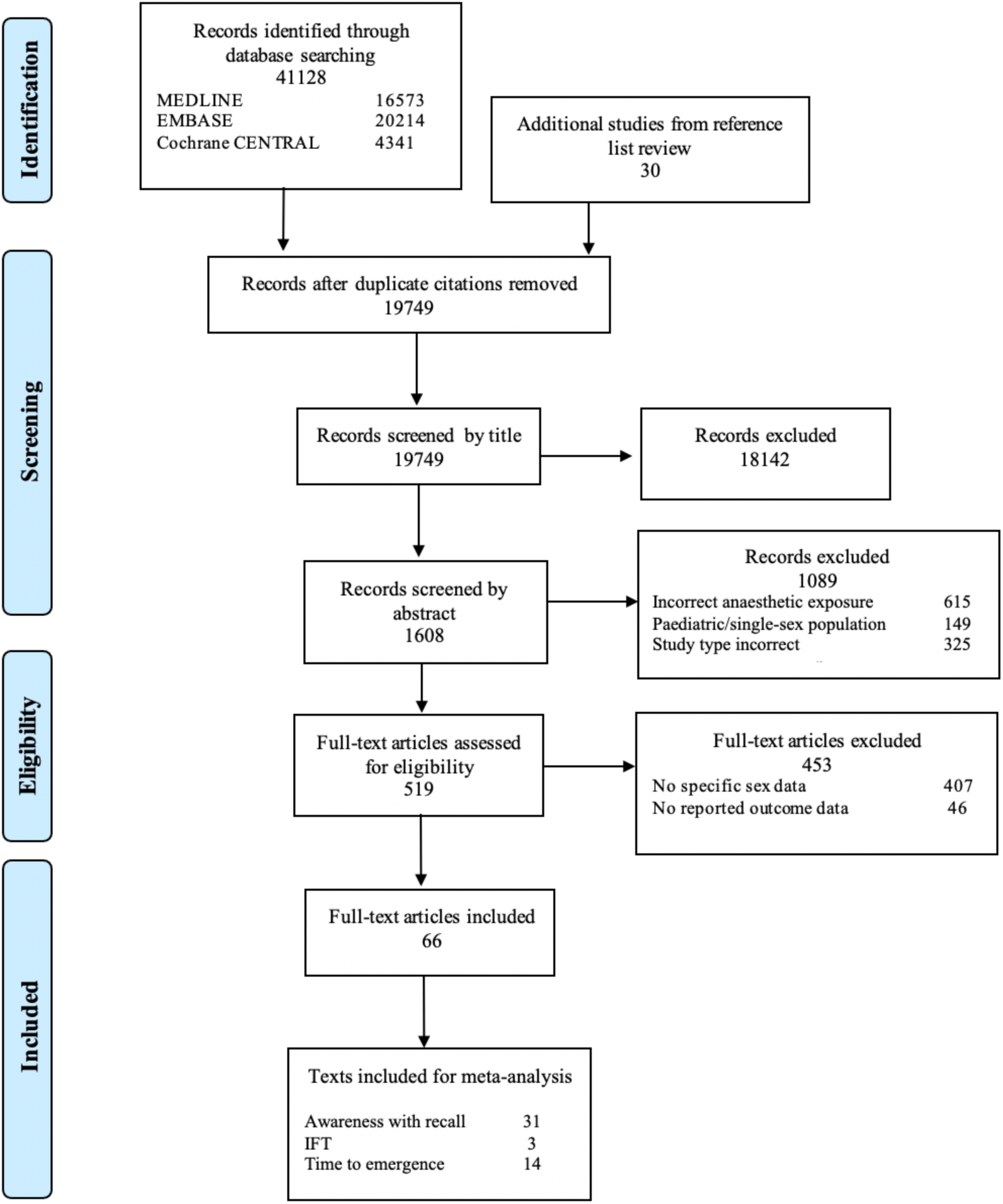
A Preferred Reporting Items for Systematic Reviews and Meta-analyses (PRISMA) flow diagram for the systematic literature review, detailing database searches, the number of abstracts and titles screened, and the number of publications identified for inclusion.

Study characteristics are outlined in table 1 (additional information in supplementary table 1). These included randomised controlled trials (RCTs) (23 of 64),^34,37-58^ non-randomised interventional trials (19 of 64),^59-77^ and cohort studies (22 of 64).^4,9,17,18,20,78-94^ Studies were conducted in 20 countries, including four multi-national studies,^9,17,18,34^ the majority of which were undertaken in Europe (22 of 64), Asia (22 of 64) and North America (11 of 64).

**Table 1.** Study design, characteristics, and outcomes reported in studies included in the systematic review.

|  | **RCTs**<br>n (%) | **Non-**<br>**randomised**<br>**interventional**<br>**trials**<br>n (%) | **Observational**<br>**studies**<br>n (%) | **Total**<br>n (%) |
| --- | --- | --- | --- | --- |
| **Total** | 23 (35.9) | 19 (29.7) | 22 (34.4) | 64 (100) |
| **Participants** |  |  |  |  |
| Females | 7385 | 3053 | 42611 | 53049 |
| Males | 9464 | 2838 | 32710 | 45012 |
| Total | 16849 | 5891 | 75321 | 98061 |
| **Location of study** |  |  |  |  |
| Africa | --- | 1 (5.2) | 1 (4.5) | 2 (3.1) |
| Asia | 9 (39.1) | 9 (47.4) | 4 (18.2) | 22 (34.4) |
| North America | 8 (34.8) | 1 (5.3) | 2 (9.1) | 11 (17.2) |
| Europe | 5 (21.7) | 8 (42.1) | 9 (41.0) | 22 (34.4) |
| Oceania | --- | -- | 3 (13.6) | 3 (4.7) |
| Multi-national | 1 (4.4) | -- | 3 (13.6) | 4 (6.2) |
| **Anaesthetic protocol** |  |  |  |  |
| Specified | 22 (95.7) | 19 (100) | -- | 41 (64.1) |
| Non-specified | 1 (4.3) | -- | 22 (100) | 23 (35.9) |
| **Main anaesthetic agents** |  |  |  |  |
| IV-based | 10 (43.5) | 14 (73.7) | 2 (9.1) | 26 (40.6) |
| Volatile-based | 8 (34.8) | 4 (21.1) | 3 (13.6) | 15 (23.4) |
| Both IV and volatile | 4 (17.4) | 1 (5.2) | -- | 5 (7.8) |
| Not specified | 1 (4.3) | -- | 17 (77.3) | 18 (28.2) |
| **Anaesthetic agents** |  |  |  |  |
| IV anaesthesia |  |  |  |  |
| Propofol | 11 (47.8) | 13 (68.4) | 2 (9.1) | 26 (40.6) |
| Thiopentone | 1 (4.3) | 2 (10.5) | 1 (4.5) | 4 (6.2) |
| Dexmedetomidine | 2 (8.7) | 1 (5.3) | -- | 3 (4.7) |
| Ketamine | 1 (4.3) | 1 (5.3) | -- | 2 (3.1) |
| Etomidate | 2 (8.7) | -- | -- | 2 (3.1) |
| Midazolam | 2 (8.7) | 4 (21.1) | -- | 6 (9.4) |
| Other | 1 (4.3) | -- | -- | 1 (1.6) |
| **Volatile anaesthesia** |  |  |  |  |
| Sevoflurane | 8 (34.8) | 4 (21.1) | 2 (9.1) | 14 (21.9) |
| Isoflurane | 5 (21.7) | 2 (10.5) | 1 (4.5) | 8 (12.5) |
| Desflurane | 4 (17.4) | 1 (5.3) | 2 (9.1) | 7 (10.9) |
| Enflurane | -- | -- | 1 (4.5) | 1 (1.6) |
| Halothane | -- | -- | 2 (9.1) | 2 (3.1) |
| Methoxyflurane | -- | -- | 1 (4.5) | 1 (1.6) |
| Ether | -- | -- | 1 (4.5) | 1 (1.6) |
| Xenon | 1 (4.3) | -- | 1 (4.5) | 2 (3.1) |
| N<sub>2</sub>O | 5 (21.7) | 2 (10.5) | 2 (9.1) | 9 (14.1) |
| **Surgical procedure** |  |  |  |  |
| Elective | 15 (65.2) | 9 (47.4) | 11 (50.0) | 35 (54.7) |
| Elective and/or emergency | 1 (4.4) | 1 (4.3) | 3 (13.6) | 5 (7.8) |
| Nil procedure | 3 (13.0) | -- | -- | 3 (4.7) |
| Not specified | 4 (17.4) | 9 (47.3) | 8 (36.4) | 21 (32.8) |
| **Anaesthetic outcome of interest** |  |  |  |  |
| Depth of anaesthesia | 7 (30.4) | 8 (42.1) | 2 (9.1) | 17 (26.6) |
| Emergence from anaesthesia | 8 (34.8) | 6 (31.6) | 3 (13.6) | 17 (26.6) |
| Awareness | 9 (39.1) | 6 (31.6) | 18 (81.8) | 33 (51.6) |
| **Abbreviations:** N<sub>2</sub>O, nitrous oxide; RCT, randomised control trial. |  |  |  |  |

Included studies had between 18 and 19 575 participants. Surgical procedures undertaken varied: the majority of studies included elective surgery (35 of 64), five studies included both elective and emergency surgery participants, two studies did not involve surgical procedures, and 21 did not specify details of the surgery.

Anaesthetic agents varied between and within studies (table 1, supplementary table 1). Forty-one of the 64 studies provided a specified anaesthetic protocol for general anaesthesia or sedation; the remainder left the choice and dosage of anaesthetic agent to the discretion of the anaesthetist. Twenty-six studies used predominantly intravenous anaesthesia, fifteen studies used predominantly volatile anaesthesia, five studies used a mixture of intravenous and volatile anaesthesia, and 18 studies did not specify anaesthetic agents used. Of the intravenous anaesthetics, 26 studies used propofol, six studies used midazolam, four studies used thiopentone, three studies dexmedetomidine, two studies each used ketamine or etomidate. Of the inhalational anaesthetics, 14 studies used sevoflurane, nine studies used nitrous oxide, eight studies used isoflurane, seven used desflurane, two studies each used halothane and xenon, and one study each used methoxyflurane, enflurane and ether.

### Risk of Bias

Of the included studies, 52 of 64 were considered good quality (fig 2). Nine studies were fair quality with some concerns for bias, the most common reasons being lack of blinding of assessors to outcome or study groups. Three studies were poor quality with high concerns for bias. Of these, none provided information on blinding of the assessor and performed post-operative awareness with recall interviews on only one occasion in the early post-operative period reducing the likelihood of capturing positive results,^45,65,73^ two did not provide demographic information on participants lost to follow-up,^65,73^ and one did not provide clear information about outcome assessment.^45^

**Figure 2.**
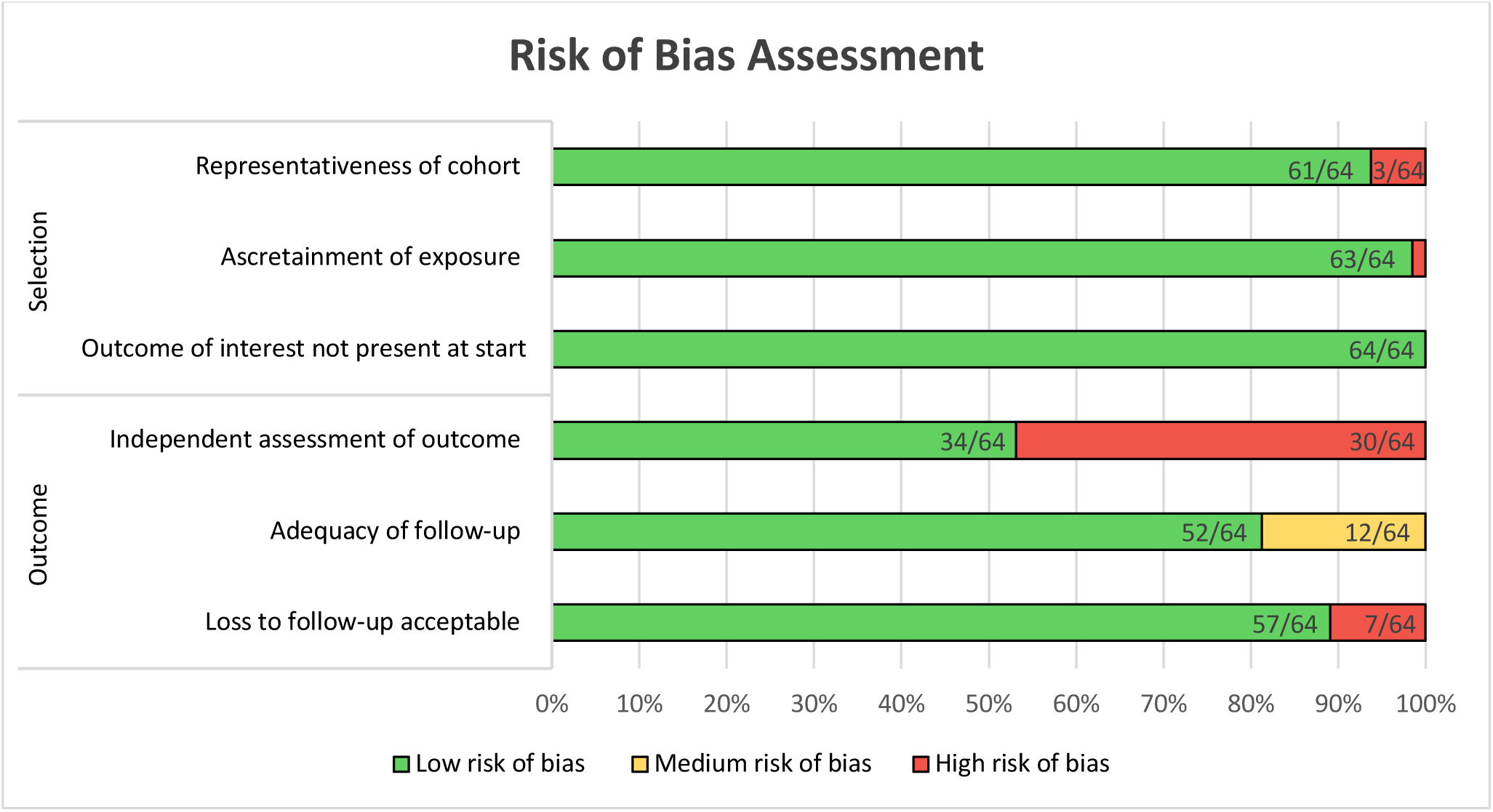
Summary of risk of bias assessment, using the Newcastle-Ottawa scale.

Studies were also reviewed using the REAPPRAISED checklist. Of the included studies, 11 studies had some concern for publication integrity (six studies on awareness with post-operative recall^37,55,65,66,81,89^; three studies on depth of anaesthesia^48,58,73^; two studies on emergence of anaesthesia^41,69^).

### Awareness with post-operative recall following general anaesthesia

Thirty-one studies (89,485 participants, 49,477 females: 40,008 males) reported awareness with post-operative recall in female and male subgroups (supplementary table 3). Confirmed awareness with post-operative recall was significantly associated with female sex (OR = 1.37, 95% CI 1.09 to 1.75; fig 3). Heterogeneity was low (p = 0.76, I_2_ = 0%). Similarly, the incidence of awareness with post-operative recall from pooled data of total (possible and confirmed) was greater in females (OR 1.38, 95% CI 1.13 to 1.68; supplementary fig 1). The observed heterogeneity was also low (p = 0.57, I_2_ = 0%).

**Figure 3.**
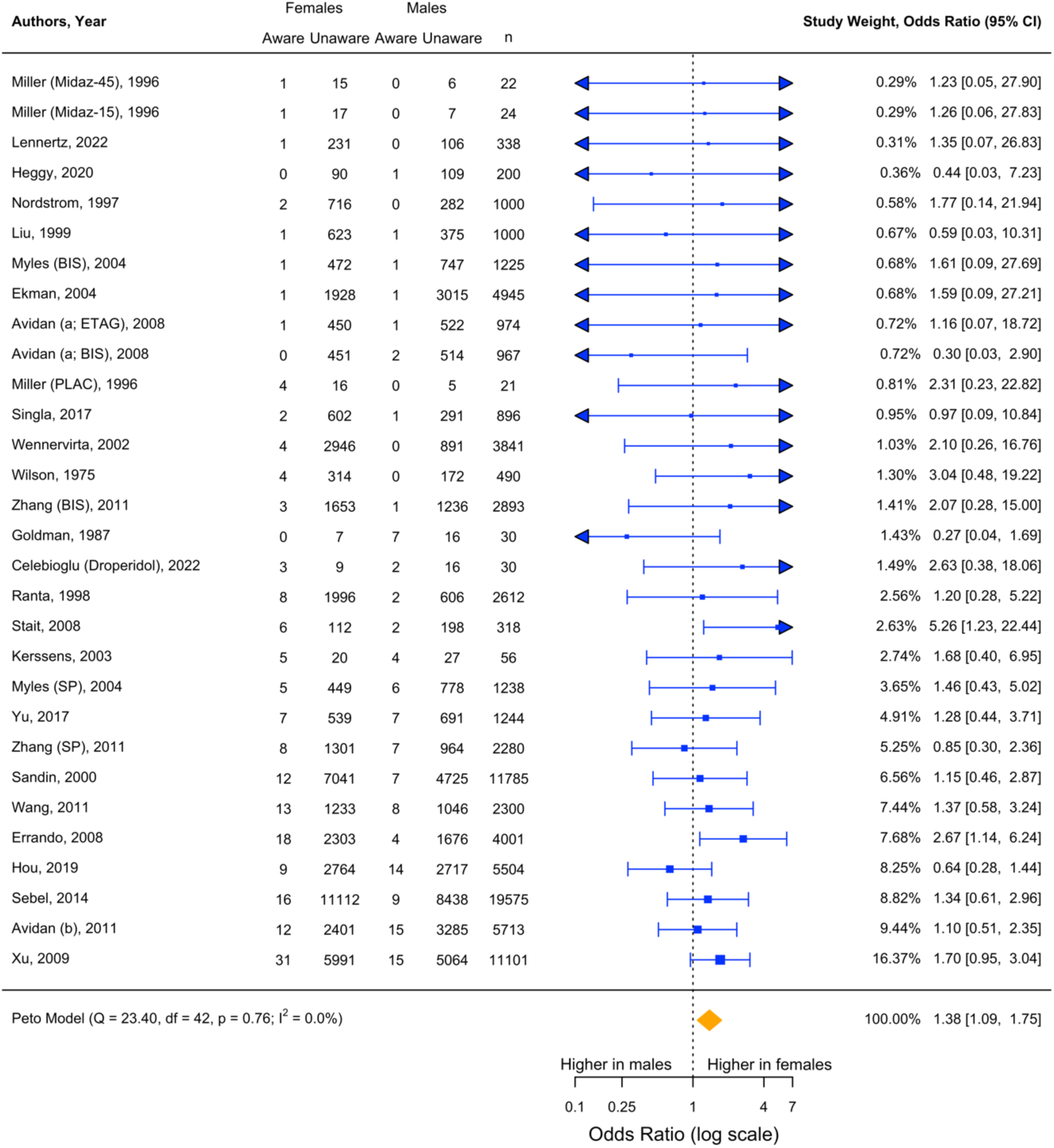
Forest plot of confirmed incidence of awareness with recall. BIS, Bispectral index; ETAG, end-tidal anaesthetic gas; Midaz-15, midazolam 15mcg/kg; Midaz-30, midazolam 30mcg/kg; Midaz-45, midazolam 45 mcg/kg; Prop, propofol; PLAC, placebo; remi, remifentanil; sevo, sevoflurane; SP, standard practice.

Both the confirmed and total incidence of awareness remained significant when using alternative models for analysis (supplementary fig 2, 3; supplementary table 4, 5) or with performance of sequential exclusion analysis (supplementary table 6, 7). When sensitivity analyses were performed on confirmed awareness with post-operative recall data following exclusion of studies with concern for publication integrity, the results were unchanged (OR 1.52, 95% CI 1.16 to 1.99; supplementary fig 4). One additional study assessed implicit memory via recall and recognition testing and found females had a 10-20% improved score compared to males (supplementary table 3).^50^

### Connected consciousness during general anaesthesia (response to command)

Three studies (618 participants, 362 female: 256 males) reported intraoperative responsiveness using the IFT (supplementary table 3).^17,18,64^ Females had a higher mean rate of connected consciousness compared to males, OR 2.09 (95% CI 1.04 to 4.23) (fig 4; heterogeneity, p = 0.80, I_2_ = 0%, τ^2^ = 0.0).

**Figure 4.**
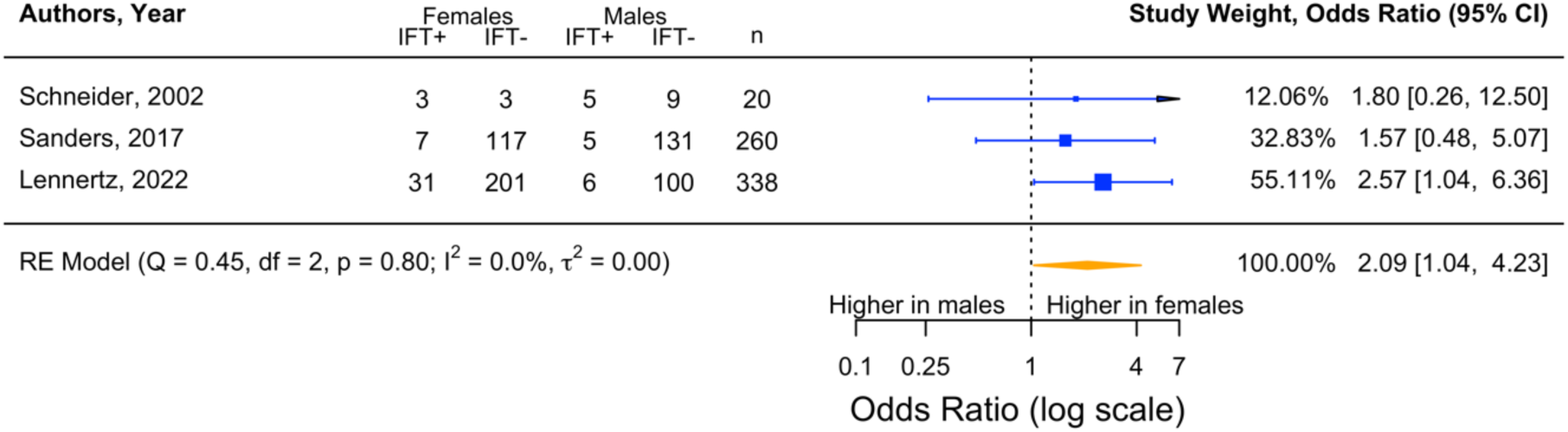
Forest plot for intra-operative response using isolated forearm technique.

### Time to intended emergence from anaesthesia

Seventeen identified studies (5399 participants, 2511 female: 2888 males; supplementary table 3, 8) reported emergence from anaesthesia by sex. All except two studies^49,54^ reported time (minute/s or second/s) to a pre-determined clinical outcome as a measure of emergence from anaesthesia. For most of these studies, the clinical outcome was eye opening (10 of 17); outcomes also included time to obeying/responding to commands (7 of 17), time to extubation (5 of 17), time to orientation (4 of 17), duration of PACU stay (3 of 17), and time to improved neurological scores (2 of 17).

Females had faster mean time to eye opening (10 studies, MD -2.28 min [95% CI -3.58 to - 0.98], fig 5). The 95% prediction interval was 6.18 min faster to 1.61 min slower in females compared to males. Heterogeneity was high (p < 0.01, I^2^ = 90.7%, τ^2^ = 2.88). Full explanation of heterogeneity investigations can be found in the appendix (supplementary table 9). When sensitivity analyses were performed on data following exclusion of studies with concern for publication integrity, the results were unchanged (OR 1.52, 95% CI 1.16 to 1.99; supplementary fig 5).

**Figure 5.**
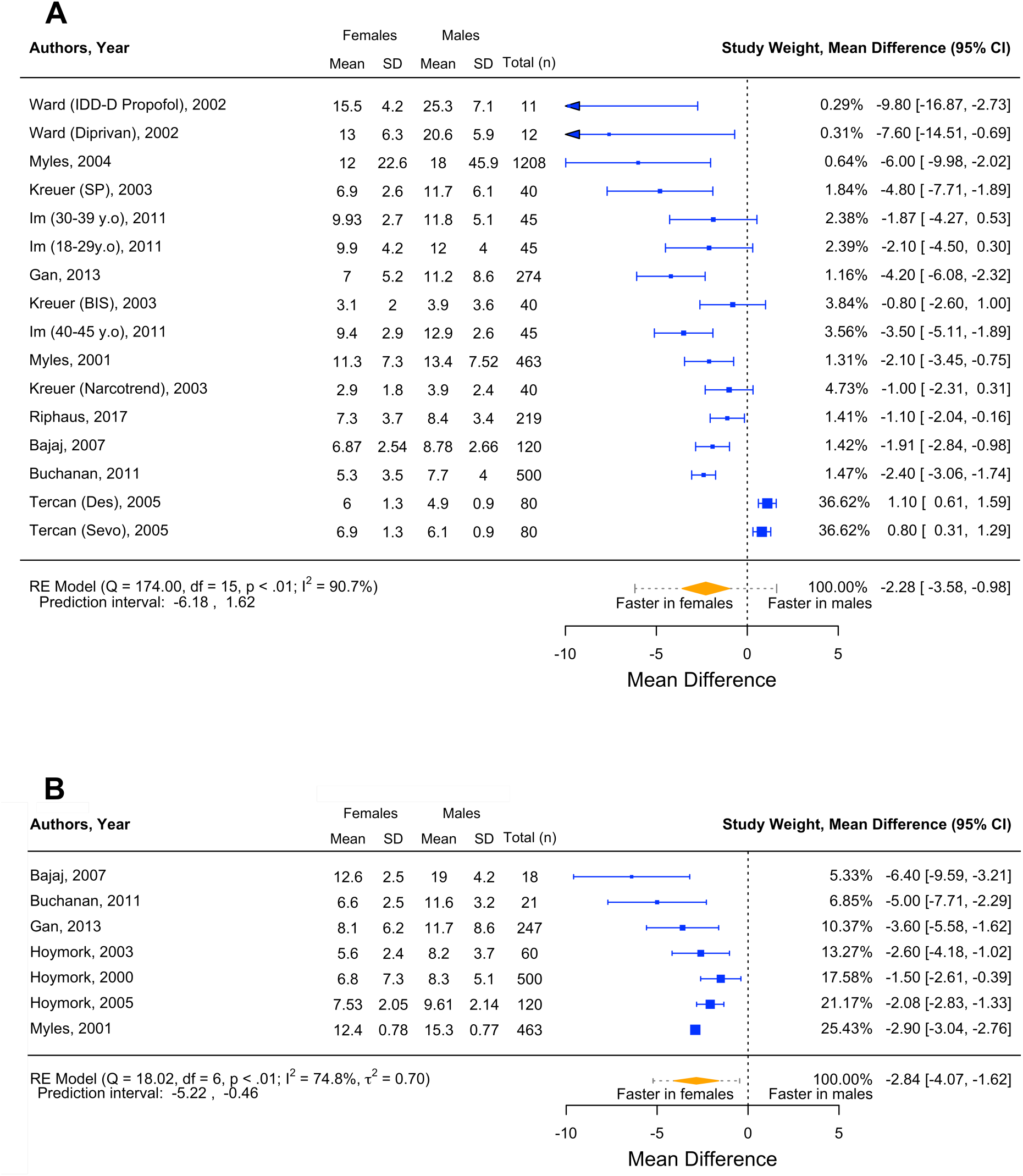
Forest plot of emergence times for (A) time to eye opening and (B) time to obeying commands in females versus males. BIS, bispectral index; Des, desflurane; Sevo, sevoflurane; SP, standard practice.

Mean time to obeying commands was faster in females (6 studies, MD -2.84 min, 95% CI - 4.07 to -1.62, 95% PI -5.22 to -0.46; fig 5, heterogeneity, p < 0.01, I^2^ = 74.8%, τ^2^ = 0.70). Time to extubation in theatre, however, did not differ significantly between females and males (4 studies; MD -0.74, 95% CI -2.58 to 1.10, 95% PI -5.44 to 3.96; supplementary fig 6). This differed from one study where patients transferred post-operatively to intensive care and median time to extubation was longer in females than in males (12 vs. 10 h; p = 0.004).^35^

The remaining studies reporting on time to orientation, duration of PACU stay and improved neurological assessment scores had conflicting results (supplementary table 3, 8). Four studies reported faster recovery time profiles for females,^34,54,69,77^ and five studies reported similar or slower recovery time profiles for females. ^40,41,49,87,94^

### Depth of Anaesthesia

Eighteen studies provided summary data on depth of anaesthesia (10290 participants, 4525 females: 5765 males). The markers of exposure included drug dosages (6 of 18), predicted effect site concentrations (EC_50_) of propofol (4 of 18), end-tidal anaesthetic agent concentrations (5 of 18), and changes in intra-operative pEEG (3 of 18). Further details of study parameters and measures of outcome are described in supplementary table 3.

Five studies reported on end-tidal agent concentrations for preventing somatic or purposeful response to noxious stimuli (4 of 5) or titration to pEEG level (1 of 5). Two studies found higher^51,62^ and three studies found lower^9,52,66^ end-tidal volatile anaesthetic concentrations in females compared to males. No study demonstrated statistically significant differences by sex.

Depth of anaesthesia as assessed by clinical presentation, including loss of response to verbal command, loss of consciousness, or sedation, was reported in nine studies. Of these, seven studies reported drug dose requirements for a given clinical indication.^38,47,48,58,61,67,75^ There was significant heterogeneity in anaesthetic drug type and dosing protocols between these studies, and accordingly results varied (supplementary table 3). The other two studies reported sedation level for given drug dose of midazolam^42^ or midazolam and ketamine.^60^ They both found that sedation levels were lighter in females than in males for their given drug protocols.

Three studies measured depth of anaesthesia by changes in processed pEEG monitoring. One study found that for similar end-tidal anaesthetic agent concentrations, females had lighter anaesthesia with a higher average intra-operative pEEG values compared to males.^36^ With propofol, the dose required to achieve deep anaesthesia was greater in females than males in one study,^90^ but not another.^63^ Lastly, one study defined adequate depth of anaesthesia as ability to successfully insert a laryngeal mask airway. Associated propofol EC_50_ levels were approximately 40% lower in females than in males.^74^

## DISCUSSION

We found female sex was associated with a greater incidence of postoperative recall and connected consciousness during intended general anaesthesia. This means females are experiencing more “failures of anaesthesia” than males. Congruent with relatively inadequate level of anaesthesia, females emerged from anaesthesia faster at the end of an operation. These data suggest that sex differences are inadequately considered as an important factor in anaesthetic titration. Due to heterogeneity of definitions for depth of anaesthesia, we were unable to quantitatively synthesize the literature but, importantly, we found no focused investigations of sex-adjusted drug dosing for the KPI of anaesthetic awareness. Standardized definitions of depth of anaesthesia, including KPIs, would be a key step in advancing anaesthesiology. Compounding this issue, we excluded 407 studies from the review due to unclear reporting of outcomes by sex. To address equity, diversity and inclusion in clinical trials, sex stratified data needs to be transparently available for core reported outcomes. Ideally these data could be provided to facilitate individual participant meta-analysis, supporting improved integration of research.

### Strengths and weaknesses of this review

This systematic review with meta-analysis comprehensively synthesizes and appraises available evidence on sex differences in anaesthetic awareness, depth and emergence. The strength of this review lies in its thorough search strategy to identify relevant literature, inclusion of prospective study designs which included all types of general anaesthetic agents across adult populations, and the performance of data extraction and literature bias assessment by two authors to reduce likelihood of errors in data analysis.

Several limitations to this review exist. Many studies were excluded from the review due to unclear sex stratification of outcomes; if these data were available for inclusion, the power of statistical analysis would have increased. For the depth of anaesthesia data, difficulties in synthesis of data from the small number of studies identified was exacerbated by heterogeneity of the included cohorts with regards to study design, definition of depth of anaesthesia and measure of anaesthetic exposure. Variation in reported outcomes and measures precluded use of meta-analyses for quantitative assessment. For the outcome of time to emergence from anaesthesia, significant between-study heterogeneity was identified. Comparatively, data on connected consciousness and recall awareness incidence were convergent.

### Possible mechanisms underpinning these findings

A growing body of evidence supports the existence of differences in both pharmacodynamic and pharmacokinetic drug profiles between females and males.^95^ Mechanisms that may contribute to this variation include differences in body composition leading to changes in volume of drug distribution.^95,96^ In addition, variations in enzymatic activity, particularly CYP450, altering drug metabolism and elimination kinetics may also contribute,^95^ although current literature to support these proposed mechanisms is conflicting.^96^ Some studies suggest important differences in pharmacokinetic profiles between females and males. For instance, females have lower plasma propofol concentrations compared to males despite the same propofol infusion scheme.^39,97^ Several mechanisms were proposed for this difference, including physiological differences in cardiac output, hepatic perfusion, and body fat composition between sexes, resulting in nontrivial differences in compartment volumes and clearance. Similarly one study identified that targeted propofol plasma concentrations for loss of consciousness in females differed when compared to measured propofol concentration; an outcome not noted for males.^67^ This discrepancy suggests a possible bias in predictive target-controlled infusion practices that may lead to systematic underdosing of females.

Our review suggests that, at least some, females have a relative insensitivity to anaesthetic agents. Some recent research has suggested that variation in sex hormones may have a modulatory role,^96,98^ with studies reporting higher end-tidal volatile requirements^13^ and higher propofol effect-site concentration requirements^16^ in pre-menopausal females during the follicular phase of their menstrual cycle. Oestrogen and progesterone have central nervous system effects,^14^ including allosteric modulation of the gamma-aminobutyric acid receptors, a major molecular target of anaesthetic action.^99^ Oestrogen may also excite the central nervous system through potentiation of the N-methyl-d-aspartate receptor and antagonizing endogenous sleep mechanisms.^100^ This provides important biological plausibility for why anaesthetic requirement may vary by sex.

Finally, the possibility of systematic underdose due to clinician practices and biases requires further exploration. Clinicians may be unaware that females will require higher mg/kg dosing than males simply to achieve the same effect site concentration. This could be compounded by dose adjustments driven by societal expectations of greater sensitivity of females to anaesthesia than males, which lacks a sound evidence base.

### Implications of findings in the clinical context and direction of future research

This review highlights an important clinical difference in response to anaesthesia between females and males that has been poorly appreciated to date. The exact aetiological basis of this apparent reduced anaesthetic sensitivity in females is unclear, mandating a coordinated scientific response to this issue that advances multiple lines of enquiry in parallel. It is plausible that this disparity stems from a complex interplay of both differences in the pharmacodynamic as well as pharmacokinetic response to the general anaesthetic agents. There are likely important implications for obstetric anaesthesia, given the high oestrogen state of pregnancy, and the high incidence of awareness with recall in this relatively understudied population. Further research into the modulatory role of sex hormones on anaesthetic dosing is also warranted. Elucidation of these mechanisms may inform future development of individualised, sex-tailored anaesthetic techniques that reduce these disparities. Most of all, we need to rule out that any practice bias effect may contribute to these outcomes and share these results widely across medicine and the public to ensure patients and carers are aware of the relevant risks.

## CONCLUSION

Our findings highlight a critical sex difference in outcomes for patients undergoing anaesthesia, with higher risk of connected consciousness during intended anaesthesia, and recall, as well as faster time to emergence in the female population. Further work needs to be done to identify underlying mechanisms of sex related differences in response to anaesthetic agents in order to better tailor anaesthesia to individual patient needs.

## Data Availability

All data produced in the present work are contained in the manuscript

## Declarations

No declarations to be made.

## Author contribution

All authors were involved in reviewing and editing of the submission. In addition, HB and RS were involved in conceptualisation, investigation, methodology and original drafting. HB, TP, ND, TM, JL, AW, and RS were involved in data curation and/or formal analysis.

## Data Sharing Statement

All data and R code are available upon request.

## Acknowledgements

The authors gratefully acknowledge the support of Prof Ian Marschner for assisting with planning of statistical analysis.

## Ethical Approval

Not applicable.

