## Supplementary Figures and Tables for "The impact of female sex on anaesthetic awareness, depth and emergence: A systematic review and meta-analysis"

**Supplementary table 1.** Further details on study design, and anaesthetic protocols.

| Author (year), country | Patient demographics | | | | Specified anaesthetic protocol | Anaesthetic agents | Elective or emergency procedure? | Anaesthetic outcome data | | |
|---|---|---|---|---|---|---|---|---|---|---|
| | Population (n) | Females (n) | Males (n) | Subgroups | | | | Depth of anaesthesia | Emergence | Awareness |
| **Randomised trials** | | | | | | | | | | |
| Arya, et al. [58] (2013), India | 70 | 35 | 35 | SP: n=35, 18♀<br>BIS: n=35, 17♀ | Yes | Induction: propofol + fentanyl | Elective | + | -- | -- |
| Avidan, et al. [57] (2011), USA | 5713 | 2413 | 3300 | BIS: n=2861, 1240 ♀<br>ETAG: n=2852, 1173♀ | Yes | Maintenance: volatiles (isoflurane, desflurane or sevoflurane) | Elective | -- | -- | + |
| Avidan, et al. [56] (2008), USA | 1941 | 902 | 1039 | BIS: n=967, 451♀<br>ETAG: n=974, 451♀ | Yes | Maintenance: volatiles (isoflurane, desflurane or sevoflurane) | Elective | -- | -- | + |
| Celebioglu, et al. [55] (2002), Turkey | 59 | 20 | 39 | Droperidol: n=30, 12♀<br>Sevoflurane: n=29, 8♀ | Yes | Induction: diazepam, etomidate, fentanyl<br>Maintenance on CPB: sevoflurane OR droperidol + fentanyl | Elective | -- | -- | + |
| Deogaonkar, et al. [54] (2011), USA | 107 | 46 | 61 | A: n=30, 14♀<br>B: n=20, 13♀<br>C: n=28, 10♀<br>D: n=29, 9♀ | Yes | Induction: thiopentone + opioid<br>Maintenance: isoflurane/$N_2O$ ± opioid | Elective | -- | + | -- |
| Gan, et al. [53] (1999), USA | 274 | 178 | 96 | Control: n=34; 16♀<br>SP: n=124; 84♀<br>BIS: n=115; 78♀ | Yes | Induction / maintenance: propofol, alfentanil<br>Maintenance: propofol + 50% $N_2O$ | Elective | -- | + | -- |
| Goto, et al. [52] (2002), Japan | 48 | 24 | 24 | Xenon 45%: n=4, 4♀<br>Xenon 50%: n=8, 4♀<br>Xenon 55%: n=8, 4♀<br>Xenon 60%: n=8, 4♀<br>Xenon 65%: n=8, 4♀<br>Xenon 70%: n=8, 4♀<br>Xenon 75%: n=4, 0♀ | Yes | Induction / maintenance: xenon | Elective | + | -- | -- |
| Greif, et al. [51] (2002), USA[b] | 20 | 10 | 10 | Placebo: n=20, 10♀<br>Auricular acupuncture: n=20, 10♀ | Yes | Induction: propofol<br>Maintenance: Desflurane, 70% $N_2O$ | Nil procedure | + | -- | -- |
| Hall, et al. [50] (2002), USA[b] | 22 | 10 | 12 | $N_2O$: 22, 10♀<br>Sevoflurane: 22, 10♀ | Yes | Sedation: sevoflurane OR $N_2O$ | Nil procedure | -- | -- | + |
| Katoh, et al. [49] (1993), Japan | 39 | 20 | 19 | Sevoflurane: n=21, 11♀<br>Isoflurane: n=18, 9♀ | Yes | Maintenance: isoflurane or sevoflurane | Elective | - | + | -- |
| Khanduja, et al. [48] (2014), India | 60 | 48 | 12 | Control: n=30, 24♀<br>Dexmedetomidine: n=30, 24♀ | Yes | Induction: thiopentone ± dexmedetomidine<br>Maintenance: isoflurane | Not specified | + | -- | -- |

| | | | | | | | | | |
|---|---|---|---|---|---|---|---|---|---|
| Kodaka, et al. [47] (2005), Japan | 78 | 40 | 38 | Sevoflurane: n=38, 20♀<br>Propofol: n=40, 20♀ | Yes | Induction: propofol or sevoflurane | Elective | + | -- | -- |
| Kreuer, et al. [46] (2003), Germany | 120 | 60 | 60 | SP: n=40, 20♀<br>BIS: n=40, 20♀<br>Nacrotrend: n=40, 20♀ | Yes | Induction / maintenance: propofol-remifentanil | Elective | -- | + | -- |
| Lallemand, et al. [45] (2003), France | 30 | 24 | 6 | Etomidate 0.2mg/kg: n=10, 9♀<br>Etomidate 0.4mg/kg: n=10, 6♀<br>Etomidate 0.6mg/kg: n=10, 9♀ | Yes | Induction: etomidate | Elective | -- | -- | + |
| Miller, et al. [44] (1996), USA | 90 | 62 | 28 | Placebo: n=21, 16♀<br>Midazolam 15mcg/kg: n=24, 17♀<br>Midazolam 30mcg/kg: n=23, 13♀<br>Midazolam 45 mcg/kg: n=22, 16♀ | Yes | Induction: midazolam, propofol, alfentanil<br>Maintenance: propofol | Elective | -- | -- | + |
| Myles, et al. [34] (2004) (multi-national) | 2463 | 927 | 1536 | BIS: n=1225, 471♀<br>SP: n=1238, 454♀ | No | Variety of anaesthetic agents | Both | -- | + | + |
| Schneider, et al. [43] (2003), Germany | 40 | 16 | 24 | Group 1: n=10, 2♀<br>Group 2: n=10, 6♀<br>Group 3: n=10, 2♀<br>Group 4: n=10, 6♀ | Yes | Induction/maintenance:<br>Group 1: Sevoflurane/low dose remifentanil<br>Group 2: Sevoflurane/high dose remifentanil<br>Group 3: Propofol/low dose remifentanil<br>Group 4: Propofol/high dose remifentanil | Elective | -- | -- | + |
| Sun, et al. [42] (2008), China | 360 | 180 | 180 | Midazolam 0.02mg/kg: 180, 90♀<br>Midazolam 0.06mg/kg: 180, 90♀ | Yes | Sedation: midazolam | Elective | + | -- | -- |
| Tercan, et al. [41] (2005), Turkey | 160 | 80 | 80 | Desflurane: n=80, 40♀<br>Sevoflurane: n=80, 40♀ | Yes | Induction: propofol, fentanyl<br>Maintenance: desflurane or sevoflurane + $N_2O$ | Elective | -- | + | -- |
| Wang, et al. [40] (2019), China | 32 | 16 | 16 | Esketamine: n=16, 8♀<br>Ketamine: n=16, 8♀ | Yes | Sedation: ketamine | Not specified | -- | + | -- |
| Ward, et al. [39] (2002), USA[b] | 12 | 6 | 6 | IDD-D: n=11, 6♀<br>Diprivan: n=12, 6♀ | Yes | Induction / maintenance: propofol | Nil procedure | -- | + | -- |

| | | | | | | | | | | |
|---|---|---|---|---|---|---|---|---|---|---|
| Xiong, et al. [38] (2019), China | 120 | 60 | 60 | Placebo: n=30, 15♀<br>Dexmedetomidine 0.4 ng/mL: n=30, 15♀<br>Dexmedetomidine 0.6 ng/mL: n=30, 15♀<br>Dexmedetomidine 0.8ng/mL: n=30, 15♀ | Yes | Induction: propofol, dexmedetomidine<br>Maintenance: propofol | Not specified | + | -- | -- |
| Zhang, et al. [37] (2011) (China) | 5173 | 2208 | 2965 | BIS: n=2919, 1237♂<br>SP: n=2309, 971♂ | Yes | Induction / maintenance: propofol | Not specified | -- | -- | + |
| **Other interventional trials** | | | | | | | | | | |
| Bajaj, et al. [76] (2007), India | 120 | 60 | 60 | -- | Yes | Induction: fentanyl, propofol, midazolam<br>Maintenance: propofol, 50% $N_2O$ | Elective | -- | + | -- |
| Bell, et al. [75] (1987), UK | 794 | 379 | 415 | -- | Yes | Sedation: midazolam | Not specified | + | -- | -- |
| Choi, et al. [74] (2016), Korea | 40 | 20 | 20 | -- | Yes | Induction: dexmedetomidine, propofol | Not specified | + | -- | -- |
| Heggy, et al. [73] (2020), Egypt | 200 | 90 | 110 | -- | Yes | Induction: propofol or thiopentone<br>Maintenance: isoflurane or sevoflurane | Elective | -- | -- | + |
| Hoymork, et al. [72] (2005), Norway | 60 | 30 | 30 | -- | Yes | Induction / maintenance: propofol | Not specified | -- | + | -- |
| Hoymork, et al. [70] (2003) (Norway) | 21 | 14 | 7 | -- | Yes | Induction / maintenance: propofol-remifentanil | Not specified | -- | + | -- |
| Hoymork, et al. [71] (2000), Norway | 18 | 9 | 9 | -- | Yes | Induction / maintenance: propofol-remifentanil | Not specified | -- | + | -- |
| Im, et al. [69] (2011), Korea | 135 | 71 | 64 | 18-29 y: n=45, 22♀<br>30-39 y: n=45, 23♀<br>40-54 y: n=45, 25♀ | Yes | Induction / maintenance: propofol-remifentanil | Elective | -- | + | -- |
| Kerssens, et al. [68] (2003), Netherlands | 56 | 25 | 31 | -- | Yes | Induction / maintenance: propofol | Elective | -- | -- | + |
| Kodaka, et al. [67] (2006), Japan[b] | 35 | 17 | 18 | | Yes | Induction / maintenance: propofol | Elective | + | -- | -- |
| Li, et al. [66] (2021), China | 390 | 333 | 57 | Low sevoflurane: n=69, 27♂<br>High sevoflurane: n=78, 30♂ | Yes | Induction/maintenance: sevoflurane, sufentanil | Elective | + | -- | + |
| Messahel, et al. [65] (2007), Saudi Arabia | 2328 | 1239 | 1089 | -- | Yes | Induction: fentanyl, thiopentone | Not specified | -- | -- | + |

| | | | | | | | | | | |
|---|---|---|---|---|---|---|---|---|---|---|
| | | | | | | Maintenance: isoflurane or sevoflurane + $N_2O$ | | | | |
| Riphaus, et al. [77] (2017), Germany | 219 | 108 | 111 | -- | Yes | Sedation: propofol | Both | -- | + | -- |
| Schneider, et al. [64] (2002), Germany | 20 | 6 | 14 | -- | Yes | Induction: alfentanil, propofol<br>Maintenance: propofol | Elective | -- | -- | + |
| Schultz, et al. [63] (2008), Germany | 20 | 10 | 10 | -- | Yes | Induction / maintenance: propofol-remifentanil | Not specified | + | -- | -- |
| Wadhwa, et al. [62] (2003), USA | 34 | 17 | 17 | -- | Yes | Maintenance: desflurane | Elective | + | -- | -- |
| Yen, et al. [61] (2011), Taiwan | 60 | 30 | 30 | -- | Yes | Sedation: midazolam, alfentanil | Elective | + | -- | -- |
| Yeo, et al. [60] (2017), Korea | 97 | 49 | 48 | -- | Yes | Sedation: midazolam, ketamine | Not specified | + | -- | -- |
| Yu, et al. [59] (2017) (China) | 1244 | 546 | 698 | -- | Yes | Maintenance:<br>CIIA group: sevoflurane + propofol<br>TIVA group: propofol | Not specified | -- | -- | + |
| **Prospective, observational studies** | | | | | | | | | | |
| Buchanan, et al. [94] (2011), Australia | 500 | 247 | 253 | -- | No | Induction: propofol or thiopentone<br>Maintenance: isoflurane, desflurane or enflurane, +/- $N_2O$ | Elective | -- | + | -- |
| Eger, et al. [9] (2003), multi-national | 4945 | 1929 | 3016 | -- | No | Maintenance: desflurane, ether, halothane, methoxyflurane, sevoflurane or xenon | Not specified | + | -- | -- |
| Ekman, et al. [93] (2004), Sweden | 4945 | 1929 | 3016 | -- | No | Variety of anaesthetic agents | Not specified | -- | -- | + |
| Errando, et al. [92] (2008), Spain | 4001 | 2321 | 1680 | -- | No | Variety of anaesthetic agents | Both | -- | -- | + |
| Goldmann, et al. [91] (1987), UK | 30 | 7 | 23 | -- | No | Induction / maintenance: fentanyl, halothane, $N_2O$ | Elective | -- | -- | + |
| Haensch, et al. [90] (2009)Germany | 656 | 417 | 239 | -- | No | Induction / maintenance: propofol | Elective | + | + | -- |
| Hou, et al. [89] (2019) (China) | 5404 | 2673 | 2731 | -- | No | Variety of anaesthetic agents | Not specified | -- | -- | + |
| Lennertz, et al. [17] (2022), multi-national | 338 | 232 | 106 | -- | No | Variety of anaesthetic agents | Not specified | -- | -- | + |

| | | | | | | | | | | |
|---|---|---|---|---|---|---|---|---|---|---|
| Liu, et al. [88] (1991), UK | 1000 | 624 | 376 | -- | No | Variety of anaesthetic agents | Not specified | -- | -- | + |
| Myles, et al. [87] (2001), Australia | 463 | 222 | 241 | -- | No | Variety of anaesthetic agents | Elective | -- | + | -- |
| Nordstrom, et al. [86] (1997), Sweden | 1000 | 718 | 282 | -- | No | Induction / maintenance: propofol, alfentanil | Not specified | -- | -- | + |
| Ranta, et al. [85] (1998), Finland | 2612 | 2004 | 608 | -- | No | Variety of anaesthetic agents | Elective | -- | -- | + |
| Sanders, et al. [18] (2017), 2017, multi-national | 260 | 124 | 136 | -- | No | Variety of anaesthetic agents | Not specified | -- | -- | + |
| Sandin, et al. [4] (2000), 2000, Sweden | 11,785 | 7053 | 4732 | | No | Variety of anaesthetic agents | Both | -- | -- | + |
| Sebel, et al. [20] (2004), USA | 19,757 | 11,310 | 8447 | -- | No | Variety of anaesthetic agents | Elective | -- | -- | + |
| Singla, et al. [84] (2017), India | 896 | 604 | 292 | -- | No | Variety of anaesthetic agents | Elective | -- | -- | + |
| Stait, et al. [83] (2008), Australia | 200 | 118 | 82 | -- | No | Variety of anaesthetic agents | Elective | -- | -- | + |
| Tamire, et al. [82] (2019), Ethiopia | 1065 | 773 | 292 | -- | No | Variety of anaesthetic agents | Both | -- | -- | + |
| Wang, et al. [81] (2011), China | 32 | 16 | 16 | -- | No | Variety of anaesthetic agents | Not specified | -- | -- | + |
| Wennervirta, et al. [80] (2002), Finland | 3841 | 2950 | 891 | -- | No | Variety of anaesthetic agents | Elective | -- | -- | + |
| Wilson, et al. [79] (1975), USA | 490 | 318 | 172 | -- | No | Variety of anaesthetic agents | Elective | -- | -- | + |
| Xu, et al. [78] (2009), China | 11,101 | 6022 | 5079 | -- | No | Variety of anaesthetic agents | Elective | -- | -- | + |

**Abbreviations**: BIS, Bispectral index; CIIA, combined intravenous and inhaled anaesthesia; CPB, cardiopulmonary bypass; dexmed, dexmedetomidine; ETAG, end-tidal anaesthetic gas concentration; RCT, randomised control trial; SP, standard practice; TIVA, total intravenous anaesthesia; UK, United Kingdom; USA, United States of America.

**+** = related data present**; --** = related data not present.

[a] Nested with data reported in Buchanan et al. (2006) and Leslie et al. (2005) papers

[b] Cross-over study or cross-over pairs within study

**Supplementary table 2.** Database Search Strategy m

| DATABASE: **Embase** Classic+Embase <1947 to 2022 August 19> | | |
|---|---|---|
| **#** | **Query** | **Results from 22 Aug 2022** |
| 1 | exp general anesthesia/ | 110,142 |
| 2 | exp propofol/ | 64,336 |
| 3 | barbiturates.mp. or exp barbituric acid derivative/ | 174,860 |
| 4 | benzodiazepines.mp. or exp benzodiazepine derivative/ | 259,708 |
| 5 | exp halothane/ or exp nitrous oxide/ or exp inhalation anesthesia/ or exp isoflurane/ or exp desflurane/ or exp sevoflurane/ or exp inhalation anesthetic agent/ or exp enflurane/ | 189,905 |
| 6 | exp ketamine/ | 51,877 |
| 7 | exp xenon/ | 9,048 |
| 8 | Adrenergic alpha-Agonists.mp. or exp alpha adrenergic receptor stimulating agent/ | 467,662 |
| 9 | exp intravenous anesthetic agent/ or exp intravenous anesthesia/ | 13,919 |
| 10 | emergence.mp. | 154,626 |
| 11 | depth of anaesthesia.mp. or exp anesthesia level/ | 8,270 |
| 12 | exp anesthetic recovery/ | 8,648 |
| 13 | exp unconsciousness/ or loss of consciousness.mp. | 97,551 |
| 14 | exp awareness/ or exp intraoperative awareness/ or isolated forearm technique.mp. [mp=title, abstract, heading word, drug trade name, original title, device manufacturer, drug manufacturer, device trade name, keyword heading word, floating subheading word, candidate term word] | 116,416 |
| 15 | 4 or 8 | 693,513 |
| 16 | 1 and 15 | 14,227 |
| 17 | 2 or 3 or 5 or 6 or 7 or 9 or 16 | 432,138 |
| 18 | 10 or 11 or 12 or 13 or 14 | 380,165 |
| 19 | 17 and 18 | 20,214 |
| DATABASE: Ovid **MEDLINE**(R) and Epub Ahead of Print, In-Process, In-Data-Review & Other Non-Indexed Citations, Daily and Versions <1946 to August 01, 2022> | | |
| **#** | **Query** | **Results from 2 Aug 2022** |
| 1 | exp Anesthesia, General/ | 61,144 |
| 2 | exp Propofol/ | 16,188 |
| 3 | exp Barbiturates/ | 54,129 |
| 4 | exp Benzodiazepines/ | 68,414 |
| 5 | exp Anesthetics, Inhalation/ | 55,870 |
| 6 | exp Xenon/ | 7,989 |
| 7 | exp Nitrous Oxide/ | 15,241 |
| 8 | exp Ketamine/ | 14,267 |
| 9 | exp Adrenergic alpha-Agonists/ | 166,202 |

| | | |
|---|---|---|
| 10 | exp Anesthetics, Intravenous/ | 84,102 |
| 11 | emergence.mp. | 129,920 |
| 12 | depth of anaesthesia.mp. | 750 |
| 13 | depth of anesthesia.mp. | 1,347 |
| 14 | eye opening.mp. | 2,160 |
| 15 | exp Orientation/ | 28,554 |
| 16 | recovery.mp. or exp Anesthesia Recovery Period/ | 555,764 |
| 17 | loss of consciousness.mp. or exp Unconsciousness/ | 48,954 |
| 18 | exp Intraoperative Awareness/ or exp Awareness/ | 21,881 |
| 19 | 1 or 2 or 3 or 4 or 5 or 6 or 7 or 8 or 9 or 10 | 413,282 |
| 20 | 11 or 14 or 15 or 16 | 711,669 |
| 21 | 12 or 13 or 17 | 50,946 |
| 22 | 18 or 20 or 21 | 779,498 |
| 23 | 19 and 22 | 22,949 |
| 24 | limit 23 to humans | 16,573 |

**Supplementary table 3.** Further individual study data on outcome measures

| Reference | Awareness parameters | Outcomes | Related variables[a] | | |
|---|---|---|---|---|---|
| | | | **Age (y)** | **Drug dosage** | **Duration of anaesthesia/ surgery (min)** |
| **Awareness** | | | | | |
| Avidan, et al. [57] (2011) | Incidence of awareness with recall (modified Brice questionnaire) | • Confirmed (n) = 27 (F:M 12:15) | BIS: 60 ± 14.2<br>ETAC: 61 ± 14.4 | Median age-adjusted MAC: 0.9 | NR |
| Avidan, et al. [56] (2008) | Incidence of awareness with recall (modified Brice questionnaire) | • Confirmed (n) = 4 (F:M 1:3)<br>• Possible (n) = 5 (F:M 2:3) | BIS: 59.5 ± 14.8<br>ETAG: 59.2 ± 14.6 | Mean time-averaged ETAG:<br>BIS: 0.8 ± 0.25 MAC<br>ETAG: 0.8 ± 0.23 MAC | NR |
| Celebioglu, et al. [55] (2002) | Incidence of awareness with recall (modified Brice questionnaire) | • Confirmed (n) = 5 (F:M 3:2) | Droperidol: 54 ± 16<br>Sevoflurane: 51 ± 15 | NR | Droperidol: 265 ± 50<br>Sevoflurane: 274 ± 69 |
| Ekman, et al. [93] (2004) | Incidence of awareness with recall (modified Brice questionnaire) | • Confirmed (n) = 2 (F:M 1:1) | SP: 49 ± 19<br>BIS: 50 ± 19 | NR | SP: 114 ± 72<br>BIS: 121 ± 72 |
| Errando, et al. [92] (2008) | Incidence of awareness with recall (modified Brice questionnaire) | • Confirmed (n) = 22 (F:M 18:4) | 51 ± 18 | NR | 120 ± 51 |
| Goldmann, et al. [91] (1987) | Incidence of awareness with recall (structured interview) | • Confirmed (n) = 7 (F:M 0:7) | 55.6 | NR | NR |
| Hall, et al. [50] (2002) | Recall/recognition scores of audio/visual prompts displayed during anaesthetic | Scores 10-20% poorer in M (LR chi-square < 0.004) | 29 ± 7 | NR | NR |
| Heggy, et al. [73] (2020) | Incidence of awareness with recall (modified Brice questionnaire) | • Confirmed (n) = 1 (F:M 0:1) | 44 | NR | NR |
| Hou, et al. [89] (2019) | Incidence of awareness with recall (modified Brice questionnaire) | • Confirmed (n) = 23 (F:M 9:14) | 52.6 ± 13.4 | NR | Mean NR |
| Kerssens, et al. [68] (2003) | Incidence of positive intra-operative response using IFT | • No gender specific data provided | 37 ± 10 | NR | Pre-surgical: 39 ± 11<br>Surgical: 45 ± 17 |
| | Incidence of awareness with recall (structured interview) | • Confirmed (n) = 9 (F:M 5:4) | | | |
| Lallemand, et al. [45] (2003) | Incidence of awareness with recall (structured interview) | • Confirmed (n) = 0 | Group 1: 52 (38-68)<br>Group 2: 51 (35-78)<br>Group 3: age 47 (32-57) | Etomidate dose<br>Group 1: 0.2mg/kg<br>Group 2: 0.4mg/kg<br>Group 3: 0.6mg/kg | NR |
| Lennertz, et al. [17] (2022) | Incidence of positive intra-operative response using IFT | • Responders: 11% (37/232; F:M 31:6) | 30 ± 6.3 | NR | NR |
| | Incidence of awareness with recall (structured interview) | • Confirmed awareness (n) = 1 (F:M 1:0) | | | |

| | | | | | |
|---|---|---|---|---|---|
| | | • Possible awareness (n) = 5 (F:M 3:2) | | | |
| Li, et al. [66] (2021) | Incidence of awareness with recall (structured interview) | • Confirmed awareness (n) = 0 | Low sevoflurane: 51.32 ± 1.1<br>High sevoflurane: 49.46 ± 1.11 | NR | Low sevoflurane: 214.14 ± 9.2<br>High sevoflurane: 2185 ± 9.0 |
| Liu, et al. [88] (1991) | Incidence of awareness with recall (structured interview) | • Confirmed (n) = 2 (F:M 1:1) | NR | NR | NR |
| Messahel, et al. [65] (2007) | Incidence of awareness with recall (structured interview) | • Confirmed (n) = 0 | 38.6 (14-104) | NR | NR |
| Miller, et al. [44] (1996) | Incidence of awareness with recall (structured interview) | • Confirmed (n) = 6 (F:M 6:0) | Placebo: 34 ± 11<br>Group 2: 35 ± 9<br>Group 3: 35 ± 9<br>Group 4: age 38 ± 12 | Propofol | Placebo: 37 ± 9<br>Group 2: 37 ± 12<br>Group 3: 40 ± 14<br>Group 4: 41 ± 15 |
| Myles, et al. [34] (2004) | Incidence of awareness with recall (modified Brice questionnaire) | • Confirmed (n) = 13 (F:M 6:7)<br>• Possible (n) = 37 (F:M 18:19) | BIS: 58.1 ± 16.5<br>SP: 57.5 ± 16.9 | NR | BIS: 192 (90-264)<br>SP: 186 (78-270) |
| Nordstrom, et al. [86] (1997) | Incidence of awareness with recall (structured interview) | • Confirmed (n) = 2 (F:M 2:0) | 42 ± 15 | NR | 56 ± 45 |
| Ranta, et al. [85] (1998) | Incidence of awareness with recall (modified Brice questionnaire) | • Confirmed (n) = 10 (F:M 8:2)<br>• Possible (n) = 9 (F:M 9:0) | NR | NR | NR |
| Sanders, et al. [18] (2017) | Incidence of positive intra-operative response using IFT | • Responders: 4.6% (12/260; F:M 7:5) | 29 ± 7 | NR | NR |
| | Incidence of awareness with recall (modified Brice questionnaire) | • Confirmed (n) = 0 | | | |
| Sandin, et al. [4] (2000) | Incidence of awareness with recall (modified Brice questionnaire) | • Confirmed (n) = 19 (F:M 12:7) | 48 ± 19 | NR | 99 ± 65 |
| Schneider, et al. [43] (2003) | Incidence of awareness with recall (modified Brice questionnaire) | • Confirmed (n) = 0 | Group 1: 35 y (22-54)<br>Group 2: 53 y (22-72)<br>Group 3: 44 y (28-66)<br>Group 4: 51 y (21-79) | NR | NR |
| Schneider, et al. [67] | Incidence of positive intra-operative response using IFT | • Responders: 40% (8/20; F:M 3:5) | Responders: 40 ± 15<br>Non-responders: 45 ± 17 | NR | NR |
| Sebel, et al. [20] (2004) | Incidence of awareness with recall (modified Brice questionnaire) | • Confirmed (n) = 25 (F:M 16:9)<br>• Possible (n) = 46 (F:M 27:19) | Responders: 52 ± 15<br>Possible responders: 48 ± 14<br>Non-responders: 48 ± 14 | NR | NR |
| Singla, et al. [84] (2017) | Incidence of awareness with recall (modified Brice | • Confirmed (n) = 3 (F:M | Mean NR | NR | NR |

| | | | | | |
|---|---|---|---|---|---|
| | questionnaire) | 2:1)<br>• Possible (n) = 4 (F:M 3:1) | <30 y 25.89%<br>30-44 y 32.48%<br>45-59 y 25.78%<br>>60 y 15.85% | | |
| Stait, et al. [83] (2008) | Incidence of awareness with recall (modified Brice questionnaire) | • Confirmed (n) = 8 (F:M 6:2) | 58 ± 14 | NR | NR |
| Tamire, et al. [82] (2019) | Incidence of awareness with recall (modified Brice questionnaire) | No significant difference | 39.39 ± 18.8 | NR | NR |
| Wang, et al. [81] (2011) | Incidence of awareness with recall (modified Brice questionnaire) | • Confirmed (n) = 21 (F:M 13:8)<br>• Possible (n) = 205 (F:M 118:87) | 45 ± 14 | NR | No awareness: 148 ± 71<br>Possible/confirmed awareness: 157 ± 82 |
| Wennervirta, et al. [80] (2002) | Incidence of awareness with recall (modified Brice questionnaire) | • Confirmed (n) = 4 (F:M 4:0)<br>• Possible (n) = 7 (F:M 7:0) | Outpatient: 37 ± 6<br>Inpatient: 46 ± 13 | NR | NR |
| Wilson, et al. [79] (1975) | Incidence of awareness with recall (modified Brice questionnaire) | • Confirmed (n) = 4 (F:M 4:0) | 44.4 (15-38) | NR | F: 98.9 (10-300)<br>M: 88.3 (20-310) |
| Xu, et al. [78] (2009) | Incidence of awareness with recall (modified Brice questionnaire) | • Confirmed (n) = 46 (F:M 31:15) | 49 ± 16 | NR | <1 h (n) =1046<br>1-2h (n) =3778<br>>2h (n) = 6089 |
| Yu, et al. [59] (2017) | Incidence of awareness with recall (modified Brice questionnaire) | • Confirmed (n) = 14 (F:M 7:7) | 53.67 ± 11.67 | NR | 96.69 ± 36.68 |
| Zhang, et al. [37] (2011) | Incidence of awareness with recall (modified Brice questionnaire) | Awareness with recall (n)<br>• Confirmed BIS group = 4 (F:M 3:1)<br>• Confirmed SP group = 15 (F:M 8:7) | BIS group: 46.95 ± 14.89<br>SP group: 46.06 ± 14.59 | NR | <1 h (n) = 867<br>1-2h (n) = 2147<br>>2h (n) = 1894 |
| **Emergence from Anaesthesia** | | | | | |
| Bajaj, et al. [76] (2007) | Time to eye opening (min) | • 6.87 ± 2.54 vs 8.78 ± 2.66 (p <0.001) | F: 41.05 ± 13.18<br>M: 40.93 ± 13.84 | Mean propofol (mg)<br>F: 721.45 ± 119.47<br>M: 737.63 ± 110.20 | F: 72.12 ± 9.82<br>M: 73.37 ± 10.3 |
| | Time to response to verbal command (min) | • 7.53 ± 2.05 vs 9.61 ± 2.14 (p <0.001) | | | |
| | Time to extubation (min) | • 8.40 ± 1.60 vs 10.79 ± 1.70 (p <0.001) | | | |
| Buchanan, et al. [94] (2011) | Time to eye opening (min) | • 5.3 ± 3.5 vs 7.7 ± 4.0 (p <0.0005) | F: 39.5 (17-75)<br>M: 39.5 (18-70) | Mean ETAC (%)<br>Desflurane: F, 4.16 ± 0.46; M, 4.53 ± 0.82<br>Sevoflurane: F, 1.43 ± 0.66; M, 1.43 ± 0.64<br>Isoflurane: F, 0.70 ± 0.33; M, 0.73 ± 0.29 | F: 70.8 ± 42.5<br>M: 65 ± 43.5 |
| | Time to obeying commands (min) | • 6.8 ± 7.3 vs 8.3 ± 5.1 (p = 0.01) | | | |
| | Duration of PACU stay (min) | • 38.8 ± 16.1 vs 33.7 ± 11.8 (p <0.001) | | | |

| | | | | | |
|---|---|---|---|---|---|
| | | | | Enflurane: F, 0.77 ± 15; M, NA | |
| Deogaonkar, et al. [54] (2011) | % 'awake' by 30mins post-op | • 83% vs 57% (p = 0.002) | Group A: 53 ± 16<br>Group B: 58 ± 15<br>Group C: 52 ± 15<br>Group D: 54 ± 16 | NR | Group A: 300 (228-366)<br>Group B: 276 (222-600)<br>Group C: 354 (294-390)<br>Group D: 306 (246-420) |
| Gan, et al. [53] (1999) | Time to eye opening | • 7.0 ± 5.2 vs 11.2 ± 8.6 (p <0.05) | F: 40 ± 13<br>M: 41 ± 15 | Propofol dose (mg/kg/min)<br>F: 0.13 ± 0.3<br>M: 0.13 ± 0.3 | F: 120 ± 63.6<br>M: 108 ± 46.2 |
| | Time to response to verbal command (min) | • 8.1 ± 6.2 vs 11.7 ± 8.6 (p <0.05) | | | |
| Haensch, et al. [90] (2009) | Time to extubation (min) | • 10.59 ± 4.90 vs 12.20 ± 5.16 (p <0.0001) | F: 53.4 ± 14.8<br>M: 50.5 ± 16.2 | NR | F: 98.3 ± 43.5<br>M: 103.7 ± 47.1 |
| Hoymork, et al. [72] (2005) | Time to response to verbal command (min) | • 5.6 ± 2.4 vs 18.2 ± 3.7 (p = 0.003) | F: 33.6 (22-47)<br>M: 36.5 (22-60) | Propofol dose (mg/kg/min)<br>F: 0.18 ± 0.03<br>M: 0.17 ± 0.03 | F: 99 ± 36<br>M: 97 ± 43 |
| Hoymork, et al. [70] (2003) | Time to response to verbal command (min) | • 6.6 ± 3.5 vs 11.6 ± 3.2 (p <0.01) | F: 36.5 (23-53)<br>M: 41.3 (25-50) | NR | 67 (41-125) |
| Hoymork, et al. [71] (2000) | Time to response to verbal command (min) | • 12.6 ± 2.5 vs 19.0 ± 4.2 (p = 0.001) | 44.8 ± 12.0 | NR | NR |
| Im, et al. [69] (2011) | Time to eye opening (min) | • 18-29 y: 9.9 ± 4.2 vs 12 ± 4.0 (NS)<br>• 30-39 y: 9.93 ± 2.7 vs 11.8 ± 5.1 (NS<br>• 40-54 y: 9.4 ± 2.9 vs 12.9 ± 2.6 (p <0.05) | Age mean NR | Propofol dose (mg)<br>18-29 y: 588.8 ± 275.9<br>30-39 y: 609.2 ± 217.5<br>40-54 y: 567.8 ± 195.9 | 18-29 y: 75.1 ± 36.4<br>30-39 y: 77.7 ± 30.8<br>40-54 y: 71.9 ± 22.4 |
| | Time to orientation (min) | • 18-29 y: 11.2 ± 4.2 vs 13.2 ± 4.1 (NS)<br>• 30-39 y: 10.9 ± 3.1 vs 13.2 ± 5.2 (NS)<br>• 40-54 y: 11.6 ± 3.7 vs 14.4 ± 2.9 (p <0.05) | | | |
| Katoh, et al. [49] (1993) | Awakening concentration (%) of volatile | • Non-significant difference | Sevoflurane: 42.9 ± 15.3<br>Isoflurane: 41.9 ±. 15.8 | -- | Sevoflurane: 145.1 ± 59.5<br>Isoflurane: 151.4 ± 40.3 |
| Kreuer, et al. [46] (2003) | Time to eye opening (min) | • SP: 6.9 ± 2.6 vs 11.7 ± 6.1 (p = 0.003) | SP: 46.1 ± 14.5<br>BIS: 43.8 ± 4.2 | Propofol dose (mg/kg/h) | SP: 108.2 ± 44.2<br>BIS: 121.2 ± 40.9 |

| | | | | | |
|---|---|---|---|---|---|
| | | • BIS: 3.1 ± 2.0 vs 3.9 ± 3.6 (NS)<br>• Nacrotrend: 2.9 ± 1.8 vs 3.9 ± 2.4 (NS) | Narcotrend: 44.8 ± 15.9 | SP: 6.8 ± 1.2<br>BIS: 4.8 ± 1.0<br>Narcotrend: 4.5 ± 1.1 | Narcotrend: 126.9 ± 67.7 |
| | Time to extubation (min) | • SP: 7.4 ± 2.6 vs 12.0 ± 6.3 (p = 0.005)<br>• BIS: 4.7 ± 3.6 vs 3.5 ± 2.0 (NS)<br>• Nacrotrend: 3.2 ± 2.2 vs 4.2 ± 2.2 (NS) | | | |
| Myles, et al. [34] (2004) | Time to eye opening[e] (min) | • 8 vs 10 (median time; p <0.001)<br>• 12 ± 22.6 vs 18 ± 45.9 (p <0.05) | BIS: 58.1 ± 16.5<br>SP: 57.5 ± 16.9 | NR | F: 162 ± 120.6<br>M: 212.4 ± 129.6 |
| | Time to extubation (for patients admitted to ICU) (h) | • 12 vs 10 (median time; p = 0.004) | | | |
| | Duration of PACU stay[f] (min) | • 59 vs 70 (median time; p = 0.004)<br>• 67.4 ± 49.1 vs 85.0 ± 56.8 (p <0.0005) | | | |
| Myles, et al. [87] (2001) | Time to eye opening (min) | • 11.3 (95% CI 10.4-12.32) vs 13.4 (95% CI 12.4 – 14.3) (p = 0.003) | F: 42 ± 15<br>M: 42 ± 28 | NR | F: 94 ± 62<br>M: 83 ± 52 |
| | Time to obeying commands (min) | • 12.4 (95% CI 11.3 – 13.7) vs 15.3 (95% CI 14.1 – 16.4) (p = 0.02) | | | |
| | Duration of PACU stay (min) | • 66 (95% CI 63 – 70) vs 64 (95% CI 60 – 67) (NS) | | | |
| Riphaus, et al. [77] (2017) | Time to eye opening (min) | • 7.3 ± 3.7 vs 8.4 ± 3.4 (p <0.01) | F: 66.9 ± 16.0<br>M: 64.8 ± 15.1 | Propofol dose (mg/kg)<br>F: 3.98 ± 1.81<br>M: 3.72 ± 1.75 | F: 23.2 ± 12.3<br>M: 23.8 ± 13.9 |
| | Time to orientation (min) | • 9.1 ± 3.9 vs 10.4 ± 3.7 (p <0.01) | | | |
| Tercan, et al. [41] (2005) | Time to eye opening (min) | • Desflurane: 6.0 ± 1.3 vs 4.9 ± 0.9 (p <0.001)<br>• Sevoflurane: 6.9 ± 1.3 vs 6.1 ± 0.9 (p = 0.001) | Desflurane:<br>F: 44.7 ± 7.6<br>M: 41.5 ± 7.5<br>Sevoflurane:<br>F: 43.5 ± 8.0<br>M: 42.7 ± 10.3 | NR | Desflurane:<br>F: 118.9 ± 13.8<br>M: 116.4 ± 9.4<br><br>Sevoflurane:<br>F: 121.6 ± 14.9<br>M: 116.2 ± 9.4 |
| | Time to extubation (min) | • Desflurane: 3.96 ± 0.6 vs 2.7 ± 0.6 (p <0.001)<br>• Sevoflurane: 4.6 ± 1.2 vs 3.97 ± 0.7 (p = 0.009) | | | |
| | Time to orientation (min) | • Desflurane: 9.7 ± 1.8 vs | | | |

| | | | | | |
|---|---|---|---|---|---|
| | | • 9.3 ± 1.4 (NS)<br>• Sevoflurane: 9.6 ± 1.8 vs 9.5 ± 1.1 (NS) | | | |
| Wang, et al. [81] (2011) | Time to recovery (OAAS/S score >5) (min) | • Esketamine: 9.5 (4-15) vs 8.5 (5-13) (NS)<br>• Racemate ketamine: 14.5 (8-17) vs 11.5 (5-26) (NS) | Esketamine: 32.0 ± 6.19<br>Ketamine: 40.0 ± 8.91 | Esketamine: 0.5 mg/kg<br>Ketamine: 1 mg/kg | NR |
| | Time to orientation (min) | • Esketamine: 11 (6-25) vs 11.5 (6-19) (NS)<br>• Racemate ketamine: 17.5 (13-21) vs 16 (6-29) (NS) | | | |
| Ward, et al. [39] (2002) | Time to eye opening (sec) | • IDD-D: 931 ± 250 vs 1517 ± 427 (p <0.01)<br>• Diprivan: 777 ± 378 vs 1234 ± 352 (p <0.01) | IDD-D: 26 ± 8.7<br>Diprivan: 26 ± 8.7 | Propofol dose (mg/kg): 8.5 | 30 ± 0 |
| **Depth of Anaesthesia** | | | | | |
| Arya, et al. [58] (2013) | Propofol dose for loss of response to verbal command (SP group) (mg/kg) | • 1.65 ± 0.44 vs 2.06 ± 0.45 (p = 0.0002) | SP: 34.2 ± 10.5<br>BIS: 32.7 ± 12.7 | -- | NR |
| | Propofol dose for sustained BIS value of 48 ± 2 (BIS group) (mg/kg) | • 1.75 ± 0.49 vs 1.83 ± 0.32 (NS) | | | |
| Bell, et al. [75] (1987) | Midazolam dose to achieve sedation (drowsy but still able to cooperate) (mg) | • 6.4 vs 7.4 (NS) | 57.5 | -- | NR |
| Choi, et al. [74] (2016) | Propofol $EC_{50}$ for successful insertion of LMA (mcg/mL) | • 3.82 ± 0.34 vs 5.46 ± 0.26 (p <0.01) | F: 35 ± 11<br>M: 35 ± 9 | Propofol mean dose<br>F: 2.77 ± 0.41 mg/kg<br>M: 3.35 ± 0.81mg/kg | NR |
| Eger, et al. [9] (2003) | MAC (%) for normalised-combined data[b] | • 1.013 ± 0.017 vs 1.005 ± 0.009 (NS) | NR | -- | NR |
| Goto, et al. [52] (2002) | Xenon ET % for 1 MAC | • 51.1 (44.6-57.6) vs 69.3 (63-75.6) | F: 71 ± 5<br>M: 70 ± 3 | -- | NR |
| Greif, et al. [51] (2002) | ET % for 1 MAC | • Control: 5.5 ± 1.0 vs 4.6 ± 0.6 (p <0.05)<br>• Intervention: 4.7 ± 0.6 vs 4.2 ± 0.6 (NS) | F: 26 ± 6<br>M: 28 ± 5 | -- | Control: 169 ± 40<br>Intervention: 171 ± 49 |
| Haensch, et al. [90] (2009) | Propofol dose during steady state anaesthesia ($D_2/E_0$ on EEG)[c] (mg/kg/ LBM/h) | • Control: 3.92 ± 0.324 vs 3.69 ± 0.230<br>• Dexmedetomidine: 2.55 ± 0.184 vs 2.76 ± 0.467 | F: 53.4 ± 14.8<br>M: 50.5 ± 16.2 | -- | F: 98.3 ± 43.5<br>M: 103.7 ± 47.1 |
| Khanduja, et al. [48] (2014) | Mean thiopentone dose for induction (loss of eyelash reflex) with or without dexmedetomidine (mg/kg) | • Control: 3.92 ± 0.324 vs 3.68 ± 0.230<br>• Dexmedetomidine: 2.55 ± 0.184 vs 2.76 ± 0.467 | Control: 40.4 ± 11.1<br>Dexmedetomidine: 42.4 ± 12.1 | -- | NR |

| | | | | | |
|---|---|---|---|---|---|
| Kodaka, et al. [67] (2006) | Measured and predicted propofol $P_{50}$ for LOC (mcg/mL) | • Predicted: 2.55 ± 0.11 vs 2.14 ± 0.10 (p <0.0001)<br>• Measured: 2.30 ± 0.28 vs 2.37 ± 0.41 (NS) | F: 40.1 ± 12.6<br>M: 36.4 ± 10.0 | -- | NR |
| Kodaka, et al. [47] (2005) | $P_{50}$ for LOC for propofol (mcg/mL) or sevoflurane (%) | • Sevoflurane: 0.92 ± 0.09 vs 0.83 ± 0.13 (NS)<br>• Propofol: 2.7 ± 0.1 vs 2.9 ± 0.2 (p <0.05) | Sevoflurane:<br>31 ± 6<br>29 ± 7<br>Propofol:<br>F: 29 ± 7<br>M: 30 ± 7 | -- | NR |
| Li, et al. [66] (2021) | Mean sevoflurane ET % for BIS 40-60 | • 1.574 ± 0.232 vs 1.637 ± 0.266 (NS) | Low sevoflurane: 51.32 ± 1.1<br>High sevoflurane: 49.46 ± 1.11 | -- | Low sevoflurane: 214.14 ± 9.2<br>High sevoflurane: 2185 ± 9.0 |
| Myles, et al. [34] (2004) | BIS value during maintenance of anaesthesia | • 46.4 ± 6.6 vs 44.6 ± 7.1 (p = 0.005) | F: 46.4 ± 18<br>M: 56.5 ± 16 | Age-adjusted MAC<br>F: 1.26 ± 0.3<br>M: 1.31 ± 0.3 | F: 34.2 ± 7.8<br>M: 34.8 ± 7.8 |
| Schultz, et al. [63] (2008) | Time to intra-operative EEG suppression (min) | • 4.78 ± 1.07 vs 4.72 ± 1.20 (NS) | F: 49.7 ± 17.7<br>M: 42.8 ± 13.5 | NR | NR |
| Sun, et al. [42] (2008) | OAA/S scores associated with midazolam dose (mg/kg) | 60-79yr:<br>• 0.02 mg/kg: 4.80 vs 4.30<br>• 0.06 mg/kg: 3.20 vs 2.80<br>40-59yr:<br>• 0.02 mg/kg: 4.76 vs 4.36<br>• 0.06 mg/kg: 3.72 vs 3.36<br>20-39yr<br>• 0.02 mg/kg: 4.76 vs 4.57<br>• 0.06 mg/kg: 4.37 vs 3.43 | Mean NR | Midazolam: 0.02mg/kg or 0.06 mg/kg | NR |
| Wadhwa, et al. [62] (2003) | ET % for 1 MAC | • 6.2 ± 0.4 vs 6.0 ± 0.3 (NS) | F: 33 ± 9<br>M: 27 ± 8 | -- | F: 24 ± 7[d]<br>M: 23 ± 10[d] |
| Xiong, et al. [38] (2019) | Propofol $EC_{50}$ for LOC (mcg/mL) | • Group 1: 2.17 vs 2.43 (p <0.05)<br>• Group 2: 1.82 vs 1.99 (p <0.05)<br>• Group 3: 1.56 vs 1.72 (p <0.05)<br>• Group 4: 1.32 vs 1.50 (p <0.05) | Group 1:<br>F: 41.8 ± 6.2<br>M: 43.3 ± 6.2<br>Group 2:<br>F: 44.3 ± 4.4<br>M: 41.1 ± 7.8<br>Group 3:<br>F: 43.5 ± 4.6<br>M: 41.7 ± 7.7<br>Group 4:<br>F: 42.5 ± 6.7<br>M: 40.1 ± 7.3 | -- | NR |
| Yen, et al. [61] (2011) | Mean midazolam (0.5mg/mL) + alfentanil (0.1mg/mL) for sedation (mL) | • 4.8 ± 0.8 vs 4.4 ± 0.7 (p<0.05) | F: 46.9 (30-65)<br>M: 49.6 (34-64) | -- | F: 11.5 ± 3.0<br>M: 11.2 ± 2.2 |
| Yeo, et al. [60] (2017) | Depth of sedation (BIS score) for midazolam 0.04mg/kg + ketamine 0.2mg/kg | • Non-significant difference | 45.5 ± 11.8 | Midazolam 0.04mg/kg and | NR |

| | |
|---|---|
| | ketamine 0.2mg/kg |

**Abbreviations**: BIS, Bispectral Index (processed EEG monitor); CI, confidence interval; $EC_{50}$, effect-site concentration for 50% patients; EEG, electroencephalography; ET, end-tidal; ETAG, end-tidal anaesthetic gas concentration; ICU, intensive care unit; IDD-D, propofol 2%; LMA, laryngeal mask airway; IFT, isolated forearm technique; LOC, loss of consciousness; ; LR, likelihood ratio; MAC, minimum alveolar concentration; NR, not reported; NS, non-significant; OAA/S, observer's assessment of alertness and sedation; P50, plasma concentration for 50% of patients; pEEG, processed electroencephalogram; PACU, post-anaesthesia care unit; SP, standard practice; TCI, target controlled infusion.

[a] Reported in mean +/- SD or mean (range) unless otherwise indicated
[b] Data reported in mean ± SEM
[c] $D2/E_0$: stage of deep anaesthesia where the raw EEG is characterised by a substantial amount of delta (0.5-3.5Hz) waves
[d] Time from induction to surgical incision/ET % measurement
[e] Study population excludes those admitted to ICU post-operatively mechanically ventilated
[f] Study population excludes those admitted directly to ICU post-operatively

**Supplementary table 4.** Summary Table of Different Models for Confirmed AAGA data.

| Statistical Model | Effect Size | | | Heterogeneity | |
|---|---|---|---|---|---|
| | **Odds Ratio Estimate** | **95% CI** | **95% prediction interval** | **P-value** | $\tau^2$ |
| **Peto** | 1.39 | 1.10 - 1.75 | N/A | 0.01 | NA |
| **Mantel-Haenszel** | 1.40 | 1.10 – 1.78 | N/A | 0.01 | NA |
| **DerSimonian-Laird*** | 1.35 | 1.13 – 1.60 | 1.13 – 1.60 | 0.00 | 0 |
| **Paule-Mandel*** | 1.35 | 1.13 – 1.60 | 1.13 – 1.60 | 0.00 | 0 |
| **Restricted Maximum Likelihood*** | 1.35 | 1.13 – 1.60 | 1.13 – 1.60 | 0.00 | 0 |
| **Restricted Maximum Likelihood with modified HKSJ** | 1.35 | 1.05 – 1.73 | 1.05-1.73 | 0.02 | 0 |
| **Restricted Maximum Likelihood without HKSJ** | 1.35 | 1.06 – 1.71 | 1.06 – 1.71 | 0.02 | 0 |
| **Generalised Linear Mixed-Effects Model†,‡** | 1.39 | 1.09 – 1.76 | 1.09 – 1.76 | 0.02 | 0 |
| **Arcsine Difference** | | | | | |
| **Restricted Maximum Likelihood with modified HKSJ** | 0.01 | 0.00 – 0.02 | 0.00 – 0.02 | 0.02 | 0 |

* Confidence interval calculated using HKSJ adjustment

† Confidence interval calculated using t-distribution

‡ Conditional model, approximate likelihood

**Supplementary table 5.** Summary Table of Different Models for Confirmed or Total AAGA data.

| Statistical Model | Effect Size | | | Heterogeneity | |
|---|---|---|---|---|---|
| | **Odds Ratio Estimate** | **95% CI** | **95% prediction interval** | **P-value** | **$\tau^2$** |
| **Peto** | 1.38 | 1.13 – 1.69 | N/A | 0.00 | NA |
| **Mantel-Haenszel** | 1.40 | 1.14 – 1.72 | N/A | 0.00 | NA |
| **DerSimonian-Laird*** | 1.35 | 1.14 – 1.59 | 1.14 – 1.59 | 0.00 | 0 |
| **Paule-Mandel*** | 1.35 | 1.14 – 1.59 | 1.14 – 1.59 | 0.00 | 0 |
| **Restricted Maximum Likelihood*** | 1.35 | 1.14 – 1.59 | 1.14 – 1.59 | 0.00 | 0 |
| **Restricted Maximum Likelihood with modified HKSJ** | 1.35 | 1.09 – 1.67 | 1.09 – 1.67 | 0.01 | 0 |
| **Restricted Maximum Likelihood without HKSJ** | 1.35 | 1.09 – 1.66 | 1.09 – 1.66 | 0.01 | 0 |
| **Generalised Linear Mixed-Effects Model†,‡** | 1.38 | 1.13 – 1.70 | 1.13 – 1.70 | 0.00 | 0 |
| **Arcsine Difference** | | | | | |
| **Restricted Maximum Likelihood*** | 0.01 | 0.00 – 0.02 | -0.01 – 0.03 | 0.01 | 0 |

* Confidence interval calculated using HKSJ adjustment

† Confidence interval calculated using t-distribution

‡ Conditional model, approximate likelihood

**Supplementary table 6**. Sequential exclusion analysis for confirmed or possible AAGA data.

| | Effect Size | | | | Heterogeneity | | |
|---|---|---|---|---|---|---|---|
| **Authors, Year** | **Odds Ratio** | **p-value** | **95% CI Lower bound** | **95% CI Upper bound** | **Cochran's Q** | **p-value** | **$I^2$ (%)** |
| **Avidan et al. (a; ETAG), 2008** | 1.39 | 0.01* | 1.10 | 1.76 | 23.85 | 0.69 | 0 |
| **Avidan et al. (a; BIS), 2008** | 1.41 | 0.00* | 1.11 | 1.78 | 21.44 | 0.81 | 0 |
| **Avidan et al. (b), 2011** | 1.42 | 0.01* | 1.11 | 1.82 | 23.46 | 0.71 | 0 |
| **Ekman et al., 2004** | 1.39 | 0.01* | 1.09 | 1.75 | 23.86 | 0.69 | 0 |
| **Errando et al., 2008** | 1.31 | 0.03* | 1.03 | 1.68 | 21.38 | 0.81 | 0 |
| **Goldman et al., 1987** | 1.43 | 0.00* | 1.13 | 1.81 | 19.95 | 0.87 | 0 |
| **Kerssens et al., 2003** | 1.38 | 0.01* | 1.09 | 1.75 | 23.80 | 0.69 | 0 |
| **Lallemand et al. (0.2mg/kg), 2003** | 1.39 | 0.01* | 1.10 | 1.75 | 23.87 | 0.74 | 0 |
| **Lallemand et al. (0.3mg/kg), 2003** | 1.39 | 0.01* | 1.10 | 1.75 | 23.87 | 0.74 | 0 |
| **Lallemand et al. (0.4mg/kg), 2003** | 1.39 | 0.01* | 1.10 | 1.75 | 23.87 | 0.74 | 0 |
| **Lennertz et al., 2022** | 1.38 | 0.01* | 1.09 | 1.75 | 23.59 | 0.70 | 0 |
| **Liu et al., 1999** | 1.39 | 0.01* | 1.10 | 1.77 | 23.52 | 0.71 | 0 |
| **Messahel et al., 2007** | 1.39 | 0.01* | 1.10 | 1.75 | 23.87 | 0.74 | 0 |
| **Miller et al. (PLAC), 1996** | 1.37 | 0.01* | 1.09 | 1.74 | 23.18 | 0.72 | 0 |
| **Miller et al. (M-15), 1996** | 1.38 | 0.01* | 1.09 | 1.75 | 23.64 | 0.70 | 0 |
| **Miller et al. (M-30), 1996** | 1.39 | 0.01* | 1.10 | 1.75 | 23.87 | 0.74 | 0 |
| **Miller et al. (M-45), 1996** | 1.38 | 0.01* | 1.09 | 1.75 | 23.65 | 0.70 | 0 |
| **Myles et al. (BIS), 2004** | 1.38 | 0.01* | 1.09 | 1.75 | 23.86 | 0.69 | 0 |
| **Myles et al. (RC), 2004** | 1.38 | 0.01* | 1.09 | 1.76 | 23.86 | 0.69 | 0 |
| **Nordstrom et al., 1997** | 1.38 | 0.01* | 1.09 | 1.74 | 23.40 | 0.71 | 0 |
| **Sanders et al., 2017** | 1.39 | 0.01* | 1.10 | 1.75 | 23.87 | 0.74 | 0 |
| **Sandin et al., 2000** | 1.40 | 0.01* | 1.10 | 1.79 | 23.69 | 0.70 | 0 |
| **Schneider et al. (Sevo, low remi), 2003** | 1.39 | 0.01* | 1.10 | 1.75 | 23.87 | 0.74 | 0 |
| **Schneider et al. (Sevo, high remi), 2003** | 1.39 | 0.01* | 1.10 | 1.75 | 23.87 | 0.74 | 0 |
| **Schneider et al. (Prop, low remi), 2003** | 1.39 | 0.01* | 1.10 | 1.75 | 23.87 | 0.74 | 0 |
| **Schneider et al. (Prop, high remi), 2003** | 1.39 | 0.01* | 1.10 | 1.75 | 23.87 | 0.74 | 0 |
| **Sebel et al., 2014** | 1.39 | 0.01* | 1.09 | 1.78 | 23.86 | 0.69 | 0 |
| **Singla et al., 2017** | 1.39 | 0.01* | 1.10 | 1.76 | 23.78 | 0.69 | 0 |
| **Wang et al., 2011** | 1.39 | 0.01* | 1.09 | 1.77 | 23.87 | 0.69 | 0 |
| **Wennervirta et al., 2002** | 1.37 | 0.01* | 1.08 | 1.74 | 23.18 | 0.72 | 0 |
| **Wilson et al., 1975** | 1.36 | 0.01* | 1.08 | 1.73 | 22.49 | 0.76 | 0 |
| **Xu et al., 2009** | 1.33 | 0.03* | 1.03 | 1.72 | 23.29 | 0.72 | 0 |
| **Yu et al., 2017** | 1.39 | 0.01* | 1.09 | 1.77 | 23.85 | 0.69 | 0 |
| **Zhang et al. (BIS),** | 1.38 | 0.01* | 1.09 | 1.75 | 23.71 | 0.70 | 0 |

| | | | | | | | |
|---|---|---|---|---|---|---|---|
| **2011** | | | | | | | |
| **Zhang et al. (RC), 2011** | 1.42 | 0.00* | 1.12 | 1.81 | 22.93 | 0.74 | 0 |
| **Celebioglu et al.,, 2022** | 1.37 | 0.01* | 1.08 | 1.73 | 22.96 | 0.73 | 0 |
| **Heggy et al., 2020** | 1.40 | 0.01* | 1.10 | 1.77 | 22.72 | 0.75 | 0 |
| **Hou et al., 2019** | 1.49 | 0.00* | 1.16 | 1.90 | 20.08 | 0.86 | 0 |
| **Li et al., 2021** | 1.39 | 0.01* | 1.10 | 1.75 | 23.87 | 0.74 | 0 |
| **Ranta et al., 1998** | 1.39 | 0.01* | 1.10 | 1.77 | 23.83 | 0.69 | 0 |
| **Stait et al., 2008** | 1.34 | 0.02* | 1.05 | 1.70 | 20.53 | 0.84 | 0 |

* **$p < 0.05$**

**Abbreviations:** BIS, Bispectral index; CI, confidence interval; ETAG, end-tidal anaesthetic gas; E-0.2, etomidate 0.2 mg/kg; E-0.3, etomidate 0.3 mg/kg; E-0.4, etomidate 0.4 mg/kg; M-15, midazolam 15 mcg/kg; M-30, midazolam 30 mcg/kg; M-45, midazolam 45 mcg/kg; PLAC, placebo; Prop, propofol; remi, remifentanil; Sevo, sevoflurane; SP, standard practice.

**Supplementary table 7.** Sequential exclusion analysis for confirmed or total AAGA data.

| | Effect Size | | | | Heterogeneity | | |
|---|---|---|---|---|---|---|---|
| **Authors, Year** | **Odds Ratio** | **p-value** | **95% CI Lower bound** | **95% CI Upper bound** | **Cochran's Q** | **p-value** | **$I^2$ (%)** |
| **Avidan et al. (a; ETAG), 2008** | 1.38 | 0.00* | 1.13 | 1.68 | 27.37 | 0.50 | 0 |
| **Avidan et al. (a; BIS), 2008** | 1.42 | 0.00* | 1.16 | 1.73 | 24.00 | 0.68 | 0 |
| **Avidan et al. (b), 2011** | 1.41 | 0.00* | 1.14 | 1.73 | 27.17 | 0.51 | 0 |
| **Ekman et al., 2004** | 1.38 | 0.00* | 1.13 | 1.69 | 27.55 | 0.49 | 0 |
| **Errando et al., 2008** | 1.33 | 0.01* | 1.08 | 1.64 | 25.11 | 0.62 | 0 |
| **Goldman et al., 1987** | 1.41 | 0.00* | 1.15 | 1.73 | 23.67 | 0.70 | 0 |
| **Kerssens et al., 2003** | 1.38 | 0.00* | 1.12 | 1.69 | 27.48 | 0.49 | 0 |
| **Lallemand et al. (0.2mg/kg), 2003** | 1.38 | 0.00* | 1.13 | 1.69 | 27.56 | 0.54 | 0 |
| **Lallemand et al. (0.3mg/kg), 2003** | 1.38 | 0.00* | 1.13 | 1.69 | 27.56 | 0.54 | 0 |
| **Lallemand et al. (0.4mg/kg), 2003** | 1.38 | 0.00* | 1.13 | 1.69 | 27.56 | 0.54 | 0 |
| **Lennertz et al., 2022** | 1.39 | 0.00* | 1.14 | 1.70 | 27.33 | 0.50 | 0 |
| **Liu et al., 1999** | 1.39 | 0.00* | 1.14 | 1.70 | 27.21 | 0.51 | 0 |
| **Messahel et al., 2007** | 1.38 | 0.00* | 1.13 | 1.69 | 27.56 | 0.54 | 0 |
| **Miller et al. (PLAC), 1996** | 1.37 | 0.00* | 1.12 | 1.68 | 26.87 | 0.53 | 0 |
| **Miller et al. (M-15), 1996** | 1.38 | 0.00* | 1.13 | 1.69 | 27.33 | 0.50 | 0 |
| **Miller et al. (M-30), 1996** | 1.38 | 0.00* | 1.13 | 1.69 | 27.56 | 0.54 | 0 |
| **Miller et al. (M-45), 1996** | 1.38 | 0.00* | 1.13 | 1.69 | 27.34 | 0.50 | 0 |
| **Myles et al. (BIS), 2004** | 1.36 | 0.00* | 1.11 | 1.67 | 27.18 | 0.51 | 0 |
| **Myles et al. (RC), 2004** | 1.38 | 0.00* | 1.12 | 1.70 | 27.55 | 0.49 | 0 |
| **Nordstrom et al., 1997** | 1.38 | 0.00* | 1.13 | 1.68 | 27.09 | 0.51 | 0 |
| **Sanders et al., 2017** | 1.38 | 0.00* | 1.13 | 1.69 | 27.56 | 0.54 | 0 |
| **Sandin et al., 2000** | 1.40 | 0.00* | 1.14 | 1.71 | 27.39 | 0.50 | 0 |
| **Schneider et al. (Sevo, low remi), 2003** | 1.38 | 0.00* | 1.13 | 1.69 | 27.56 | 0.54 | 0 |
| **Schneider et al. (Sevo, high remi), 2003** | 1.38 | 0.00* | 1.13 | 1.69 | 27.56 | 0.54 | 0 |
| **Schneider et al. (Prop, low remi), 2003** | 1.38 | 0.00* | 1.13 | 1.69 | 27.56 | 0.54 | 0 |
| **Schneider et al. (Prop, high remi), 2003** | 1.38 | 0.00* | 1.13 | 1.69 | 27.56 | 0.54 | 0 |
| **Sebel et al., 2014** | 1.44 | 0.00* | 1.15 | 1.79 | 26.93 | 0.52 | 0 |
| **Singla et al., 2017** | 1.39 | 0.00* | 1.13 | 1.70 | 27.53 | 0.49 | 0 |
| **Wang et al., 2011** | 1.38 | 0.00* | 1.13 | 1.70 | 27.56 | 0.49 | 0 |
| **Wennervirta et al., 2002** | 1.35 | 0.00* | 1.11 | 1.66 | 25.63 | 0.59 | 0 |
| **Wilson et al., 1975** | 1.37 | 0.00* | 1.12 | 1.67 | 26.18 | 0.56 | 0 |
| **Xu et al., 2009** | 1.34 | 0.01* | 1.09 | 1.67 | 27.00 | 0.52 | 0 |
| **Yu et al., 2017** | 1.39 | 0.00* | 1.13 | 1.70 | 27.54 | 0.49 | 0 |
| **Zhang et al. (BIS), 2011** | 1.38 | 0.00* | 1.13 | 1.68 | 27.40 | 0.50 | 0 |

| | | | | | | | |
|---|---|---|---|---|---|---|---|
| **Zhang et al. (RC), 2011** | 1.41 | 0.00* | 1.15 | 1.73 | 26.64 | 0.54 | 0 |
| **Celebioglu et al.,, 2022** | 1.37 | 0.00* | 1.12 | 1.67 | 26.65 | 0.54 | 0 |
| **Heggy et al., 2020** | 1.39 | 0.00* | 1.14 | 1.70 | 26.42 | 0.55 | 0 |
| **Hou et al., 2019** | 1.45 | 0.00* | 1.18 | 1.79 | 23.89 | 0.69 | 0 |
| **Li et al., 2021** | 1.38 | 0.00* | 1.13 | 1.69 | 27.56 | 0.54 | 0 |
| **Ranta et al., 1998** | 1.36 | 0.00* | 1.11 | 1.67 | 27.01 | 0.52 | 0 |
| **Stait et al., 2008** | 1.35 | 0.00* | 1.10 | 1.65 | 24.23 | 0.67 | 0 |

*** $p < 0.05$**

**Abbreviations:** BIS, Bispectral index; CI, confidence interval; ETAG, end-tidal anaesthetic gas; E-0.2, etomidate 0.2 mg/kg; E-0.3, etomidate 0.3 mg/kg; E-0.4, etomidate 0.4 mg/kg; M-15, midazolam 15 mcg/kg; M-30, midazolam 30 mcg/kg; M-45, midazolam 45 mcg/kg; PLAC, placebo; Prop, propofol; remi, remifentanil; Sevo, sevoflurane; SP, standard practice.

**Supplementary table 8.** Studies reporting data on emergence from anesthesia

| Anesthetic parameter | Studies (n) | Population (n) | | | Measure | Outcome[a] | |
|---|---|---|---|---|---|---|---|
| | | Female | Male | Total | | | |
| **Eye opening** | 10 | 1959 | 2507 | 4466 | Time (minutes) | Range across studies<br>Females<br>Males | <br>2.9 – 15.5<br>3.9 – 25.3 |
| **Response to/obeying verbal command** | 7 | 769 | 696 | 1456 | Time (minutes) | Range across studies<br>Females<br>Males | <br>5.6 – 12.4<br>8.3 – 19.0 |
| **Extubation** | 5 | 1544 | 1975 | 3519 | Time (minutes) | Range across studies[b]<br>Females<br>Males | <br>3.2 – 10.6<br>3.5 – 12.2 |
| **Duration of PACU stay** | 3 | 1396 | 2030 | 3426 | Time (minutes) | Range across studies<br>Females<br>Males | <br>38.8 – 67.4<br>33.7 – 64.0 |
| **Orientation** | 4 | 275 | 271 | 546 | Time (minutes) | Range across studies<br>Females<br>Males | <br>9.1 – 17.5<br>9.3 - 16 |
| **OAA/S score >5** | 1 | 16 | 16 | 32 | Time (minutes) | Females<br>Males | 9.5 – 14.5<br>9.3 – 9.5 |
| **Other** | | | | | | | |
| % 'awake' by 30 minutes | 1 | 46 | 61 | 107 | % patients | Female<br>Male | 83%<br>57% |
| Awakening concentration (%) of volatile | 1 | 20 | 19 | 39 | % volatile | No significant difference | |

[a]Further specifications of outcomes for each study are available in supplementary material
[b]Excluding study that reported time to extubation for patients admitted to ICU[34]
**Abbreviations:** OAA/S: observer's assessment of alertness and sedation.

**Supplementary table 9.** Sequential exclusion analysis for time to eye opening emergence data.

| Author, year | Effect size | | | | Heterogeneity | | | |
|---|---|---|---|---|---|---|---|---|
| | Mean difference (min)[1] | p-value | 95% CI lower bound | 95% CI upper bound | Cochran's Q | p-value | Tau² | I² (%) |
| **Buchanan et al. (b), 2011** | -2.08 | 0.00$^{\dagger}$ | -3.39 | -0.77 | 135.88 | 0 | 3.44 | 90.76 |
| **Buchanan et al. (a), 2006** | -1.89 | 0.01$^{\dagger}$ | -3.12 | -0.66 | 158.76 | 0 | 2.81 | 90.58 |
| **Gan et al., 2013** | -1.90 | 0.00$^{\dagger}$ | -3.10 | -0.69 | 165.47 | 0 | 2.80 | 90.67 |
| **Kreuer et al. (SP), 2003** | -2.19 | 0.00$^{\dagger}$ | -3.47 | -0.91 | 173.88 | 0 | 3.35 | 91.95 |
| **Kreuer et al. (BIS), 2003** | -2.19 | 0.00$^{\dagger}$ | -3.48 | -0.90 | 173.39 | 0 | 3.40 | 91.87 |
| **Kreuer et al. (Narcotrend), 2003** | -2.19 | 0.00$^{\dagger}$ | -3.49 | -0.89 | 172.24 | 0 | 3.43 | 91.55 |
| **Riaphus et al., 2017** | -2.26 | 0.00$^{\dagger}$ | -3.39 | -1.14 | 119.10 | 0 | 2.22 | 84.78 |
| **Tercan et al. (Des), 2005** | -2.27 | 0.00$^{\dagger}$ | -3.43 | -1.11 | 137.91 | 0 | 2.49 | 86.19 |
| **Tercan et al. (Sevo), 2005** | -1.91 | 0.00$^{\dagger}$ | -3.02 | -0.81 | 167.33 | 0 | 2.76 | 90.64 |
| **Ward et al. (IDD-D Propofol), 2002** | -1.96 | 0.00$^{\dagger}$ | -3.13 | -0.78 | 169.92 | 0 | 2.90 | 91.04 |
| **Ward et al. (Diprivan), 2002** | -2.13 | 0.00$^{\dagger}$ | -3.44 | -0.81 | 164.28 | 0 | 3.48 | 91.66 |
| **Bajaj et al., 2007** | -2.10 | 0.00$^{\dagger}$ | -3.39 | -0.80 | 172.24 | 0 | 3.36 | 92.07 |
| **Im et al. (18-29y.o), 2011** | -2.11 | 0.00$^{\dagger}$ | -3.41 | -0.82 | 172.71 | 0 | 3.37 | 92.09 |
| **Im et al. (30-39 y.o), 2011** | -1.96 | 0.01$^{\dagger}$ | -3.23 | -0.69 | 160.18 | 0 | 3.09 | 91.28 |
| **Im et al. (40-45 y.o), 2011** | -1.89 | 0.00$^{\dagger}$ | -3.06 | -0.72 | 166.61 | 0 | 2.73 | 90.49 |
| **Myles et al., 2004** | -2.11 | 0.00$^{\dagger}$ | -3.41 | -0.80 | 168.31 | 0 | 3.44 | 92.00 |
| **Myles et al., 2001** | -2.15 | <0.05 | -3.35 | -0.94 | 179.39 | 0 | 3.20 | 91.23 |

[1]Females - Males

| Authors, Year | Females Aware | Females Unaware | Males Aware | Males Unaware | n | Study Weight | Odds Ratio (95% CI) |
|---|---|---|---|---|---|---|---|
| Miller (Midaz-45), 1996 | 1 | 15 | 0 | 6 | 22 | 0.21% | 1.23 [0.05, 27.90] |
| Miller (Midaz-15), 1996 | 1 | 17 | 0 | 7 | 24 | 0.21% | 1.26 [0.06, 27.83] |
| Heggy, 2020 | 0 | 90 | 1 | 109 | 200 | 0.26% | 0.44 [0.03, 7.23] |
| Nordstrom, 1997 | 2 | 716 | 0 | 282 | 1000 | 0.43% | 1.77 [0.14, 21.94] |
| Liu, 1999 | 1 | 623 | 1 | 375 | 1000 | 0.49% | 0.59 [0.03, 10.31] |
| Ekman, 2004 | 1 | 1928 | 1 | 3015 | 4945 | 0.50% | 1.59 [0.09, 27.21] |
| Miller (PLAC), 1996 | 4 | 16 | 0 | 5 | 21 | 0.59% | 2.31 [0.23, 22.82] |
| Avidan (a; ETAG), 2008 | 2 | 449 | 1 | 522 | 974 | 0.78% | 2.27 [0.23, 22.03] |
| Wilson, 1975 | 4 | 314 | 0 | 172 | 490 | 0.95% | 3.04 [0.48, 19.22] |
| Zhang (BIS), 2011 | 3 | 1653 | 1 | 1236 | 2893 | 1.03% | 2.07 [0.28, 15.00] |
| Goldman, 1987 | 0 | 7 | 7 | 16 | 30 | 1.04% | 0.27 [0.04, 1.69] |
| Celebioglu (Droperidol), 2022 | 3 | 9 | 2 | 16 | 30 | 1.09% | 2.63 [0.38, 18.06] |
| Lennertz, 2022 | 4 | 228 | 2 | 104 | 338 | 1.34% | 0.91 [0.16, 5.18] |
| Avidan (a; BIS), 2008 | 1 | 450 | 5 | 511 | 967 | 1.56% | 0.30 [0.06, 1.49] |
| Singla, 2017 | 5 | 599 | 2 | 290 | 896 | 1.61% | 1.20 [0.25, 5.87] |
| Stait, 2008 | 6 | 112 | 2 | 198 | 318 | 1.92% | 5.26 [1.23, 22.44] |
| Kerssens, 2003 | 5 | 20 | 4 | 27 | 56 | 2.00% | 1.68 [0.40, 6.95] |
| Wennervirta, 2002 | 11 | 2939 | 0 | 891 | 3841 | 2.05% | 2.92 [0.76, 11.17] |
| Ranta, 1998 | 17 | 1987 | 2 | 606 | 2612 | 3.54% | 2.05 [0.71, 5.97] |
| Yu, 2017 | 7 | 539 | 7 | 691 | 1244 | 3.59% | 1.28 [0.44, 3.71] |
| Zhang (SP), 2011 | 8 | 1301 | 7 | 964 | 2280 | 3.83% | 0.85 [0.30, 2.36] |
| Sandin, 2000 | 12 | 7041 | 7 | 4725 | 11785 | 4.79% | 1.15 [0.46, 2.87] |
| Wang, 2011 | 13 | 1233 | 8 | 1046 | 2300 | 5.43% | 1.37 [0.58, 3.24] |
| Errando, 2008 | 18 | 2303 | 4 | 1676 | 4001 | 5.60% | 2.67 [1.14, 6.24] |
| Myles (BIS), 2004 | 12 | 461 | 11 | 741 | 1225 | 5.63% | 1.79 [0.77, 4.18] |
| Hou, 2019 | 9 | 2764 | 14 | 2717 | 5504 | 6.02% | 0.64 [0.28, 1.44] |
| Myles (SP), 2004 | 12 | 442 | 15 | 769 | 1238 | 6.45% | 1.41 [0.64, 3.10] |
| Avidan (b), 2011 | 12 | 2401 | 15 | 3285 | 5713 | 6.89% | 1.10 [0.51, 2.35] |
| Xu, 2009 | 31 | 5991 | 15 | 5064 | 11101 | 11.95% | 1.70 [0.95, 3.04] |
| Sebel, 2014 | 43 | 11085 | 28 | 8419 | 19575 | 18.24% | 1.16 [0.73, 1.86] |
| Peto Model (Q = 27.09, df = 42, p = 0.57; $I^2$ = 0.0%) | | | | | | 100.00% | 1.38 [1.13, 1.68] |

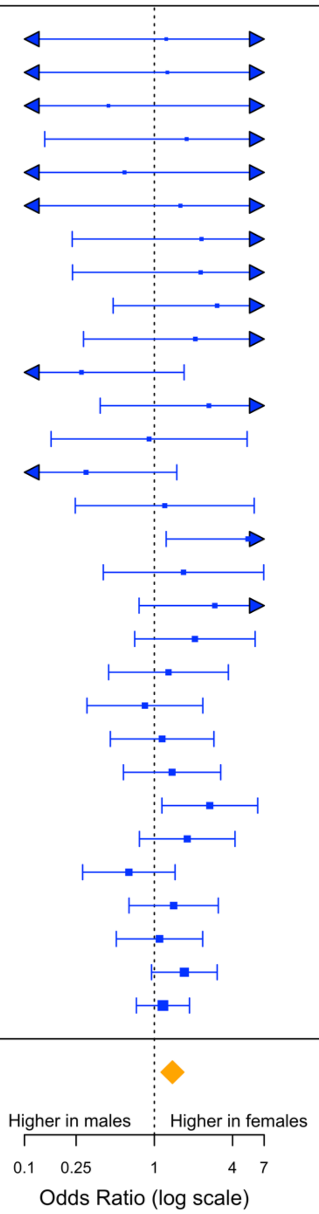


**Supplementary figure 1.** Forest plot of total (possible + confirmed) incidence of awareness with recall. BIS, Bispectral index; ETAG, end-tidal anaesthetic gas; Midaz-15, midazolam 15mcg/kg; Midaz-30, midazolam 30mcg/kg; Midaz-45, midazolam 45 mcg/kg; Prop, propofol; PLAC, placebo; remi, remifentanil; sevo, sevoflurane; SP, standard practice.

| Authors, Year | Females Aware | Females Unaware | Males Aware | Males Unaware | n | Odds Ratio [95% CI] |
|---|---|---|---|---|---|---|
| Wilson, 1975 | 4 | 314 | 0 | 172 | 490 | 3.04 [0.48, 19.22] |
| Goldman, 1987 | 0 | 7 | 7 | 16 | 30 | 0.27 [0.04, 1.69] |
| Miller (PLAC), 1996 | 4 | 16 | 0 | 5 | 21 | 2.31 [0.23, 22.82] |
| Miller (Midaz-15), 1996 | 1 | 17 | 0 | 7 | 24 | 1.26 [0.06, 27.83] |
| Miller (Midaz-45), 1996 | 1 | 15 | 0 | 6 | 22 | 1.23 [0.05, 27.90] |
| Nordstrom, 1997 | 2 | 716 | 0 | 282 | 1000 | 1.77 [0.14, 21.94] |
| Ranta, 1998 | 8 | 1996 | 2 | 606 | 2612 | 1.20 [0.28, 5.22] |
| Liu, 1999 | 1 | 623 | 1 | 375 | 1000 | 0.59 [0.03, 10.31] |
| Sandin, 2000 | 12 | 7041 | 7 | 4725 | 11785 | 1.15 [0.46, 2.87] |
| Wennervirta, 2002 | 4 | 2946 | 0 | 891 | 3841 | 2.10 [0.26, 16.76] |
| Kerssens, 2003 | 5 | 20 | 4 | 27 | 56 | 1.68 [0.40, 6.95] |
| Ekman, 2004 | 1 | 1928 | 1 | 3015 | 4945 | 1.59 [0.09, 27.21] |
| Myles (BIS), 2004 | 1 | 472 | 1 | 747 | 1225 | 1.61 [0.09, 27.69] |
| Myles (SP), 2004 | 5 | 449 | 6 | 778 | 1238 | 1.46 [0.43, 5.02] |
| Avidan (a; ETAG), 2008 | 1 | 450 | 1 | 522 | 974 | 1.16 [0.07, 18.72] |
| Avidan (a; BIS), 2008 | 0 | 451 | 2 | 514 | 967 | 0.30 [0.03, 2.90] |
| Errando, 2008 | 18 | 2303 | 4 | 1676 | 4001 | 2.67 [1.14, 6.24] |
| Stait, 2008 | 6 | 112 | 2 | 198 | 318 | 5.26 [1.23, 22.44] |
| Xu, 2009 | 31 | 5991 | 15 | 5064 | 11101 | 1.70 [0.95, 3.04] |
| Avidan (b), 2011 | 12 | 2401 | 15 | 3285 | 5713 | 1.10 [0.51, 2.35] |
| Wang, 2011 | 13 | 1233 | 8 | 1046 | 2300 | 1.37 [0.58, 3.24] |
| Zhang (BIS), 2011 | 3 | 1653 | 1 | 1236 | 2893 | 2.07 [0.28, 15.00] |
| Zhang (SP), 2011 | 8 | 1301 | 7 | 964 | 2280 | 0.85 [0.30, 2.36] |
| Sebel, 2014 | 16 | 11112 | 9 | 8438 | 19575 | 1.34 [0.61, 2.96] |
| Singla, 2017 | 2 | 602 | 1 | 291 | 896 | 0.97 [0.09, 10.84] |
| Yu, 2017 | 7 | 539 | 7 | 691 | 1244 | 1.28 [0.44, 3.71] |
| Hou, 2019 | 9 | 2764 | 14 | 2717 | 5504 | 0.64 [0.28, 1.44] |
| Heggy, 2020 | 0 | 90 | 1 | 109 | 200 | 0.44 [0.03, 7.23] |
| Lennertz, 2022 | 1 | 231 | 0 | 106 | 338 | 1.35 [0.07, 26.83] |
| Celebioglu (Droperidol), 2022 | 3 | 9 | 2 | 16 | 30 | 2.63 [0.38, 18.06] |
| Peto Model (Q = 23.40, df = 42, p = 0.76; $I^2$ = 0.0%) | | | | | | 1.38 [1.09, 1.75] |
| MH Method (Q = 16.92, df = 42, p = 0.96; $I^2$ = 0.0%) | | | | | | 1.39 [1.09, 1.77] |
| DL Method (Q = 18.92, df = 42, p = 1.00; $I^2$ = 0.0%, $\tau^2$ = 0.00) | | | | | | 1.35 [1.14, 1.59] |
| PM Method (Q = 18.92, df = 42, p = 1.00; $I^2$ = 0.0%, $\tau^2$ = 0.00) | | | | | | 1.35 [1.14, 1.59] |
| REML Method (Q = 18.92, df = 42, p = 1.00; $I^2$ = 0.0%, $\tau^2$ = 0.00) | | | | | | 1.35 [1.14, 1.59] |
| REML Method, ad hoc SE adj. (Q = 18.92, df = 42, p = 1.00; $I^2$ = 0.0%, $\tau^2$ = 0.00) | | | | | | 1.35 [1.05, 1.72] |
| REML Method, z-distribution SE (Q = 18.92, df = 42, p = 1.00; $I^2$ = 0.0%, $\tau^2$ = 0.00) | | | | | | 1.35 [1.06, 1.71] |
| GLMM (CM, AL) (Q (LRT) = 29.94, df = 29, p = 0.42; $I^2$ = 0.0%, $\tau^2$ = 0.00) | | | | | | 1.38 [1.09, 1.75] |

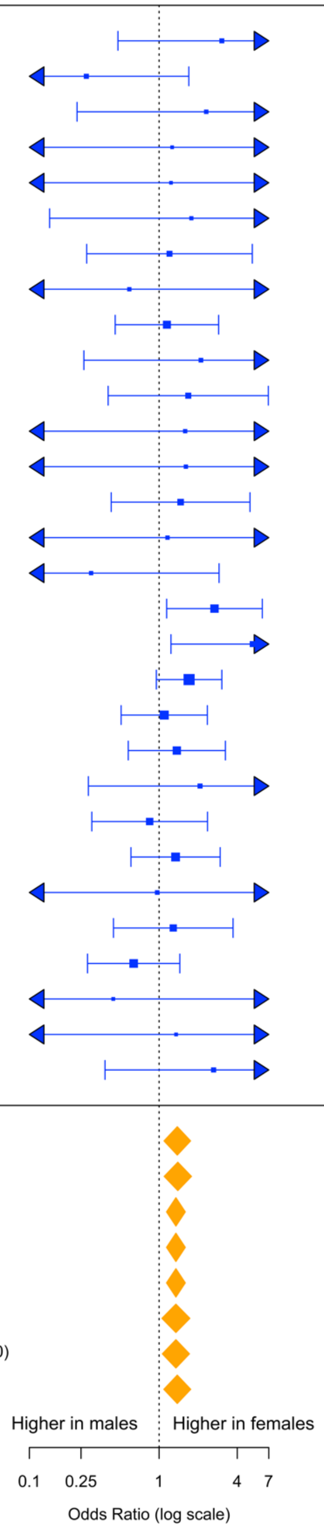


**Supplementary figure 2.** Summary forest plot of confirmed awareness data for differing methods of analysis. BIS, Bispectral index; ETAG, end-tidal anaesthetic gas; Midaz-15, midazolam 15mcg/kg; Midaz-30, midazolam 30mcg/kg; Midaz-45, midazolam 45 mcg/kg; Prop, propofol; PLAC, placebo; remi, remifentanil; sevo, sevoflurane; SP, standard practice.

| Authors, Year | Females Aware | Females Unaware | Males Aware | Males Unaware | n | Odds Ratio [95% CI] |
|---|---|---|---|---|---|---|
| Wilson, 1975 | 4 | 314 | 0 | 172 | 490 | 3.04 [0.48, 19.22] |
| Goldman, 1987 | 0 | 7 | 7 | 16 | 30 | 0.27 [0.04, 1.69] |
| Miller (PLAC), 1996 | 4 | 16 | 0 | 5 | 21 | 2.31 [0.23, 22.82] |
| Miller (Midaz-15), 1996 | 1 | 17 | 0 | 7 | 24 | 1.26 [0.06, 27.83] |
| Miller (Midaz-45), 1996 | 1 | 15 | 0 | 6 | 22 | 1.23 [0.05, 27.90] |
| Nordstrom, 1997 | 2 | 716 | 0 | 282 | 1000 | 1.77 [0.14, 21.94] |
| Ranta, 1998 | 17 | 1987 | 2 | 606 | 2612 | 2.05 [0.71, 5.97] |
| Liu, 1999 | 1 | 623 | 1 | 375 | 1000 | 0.59 [0.03, 10.31] |
| Sandin, 2000 | 12 | 7041 | 7 | 4725 | 11785 | 1.15 [0.46, 2.87] |
| Wennervirta, 2002 | 11 | 2939 | 0 | 891 | 3841 | 2.92 [0.76, 11.17] |
| Kerssens, 2003 | 5 | 20 | 4 | 27 | 56 | 1.68 [0.40, 6.95] |
| Ekman, 2004 | 1 | 1928 | 1 | 3015 | 4945 | 1.59 [0.09, 27.21] |
| Myles (BIS), 2004 | 12 | 461 | 11 | 741 | 1225 | 1.79 [0.77, 4.18] |
| Myles (SP), 2004 | 12 | 442 | 15 | 769 | 1238 | 1.41 [0.64, 3.10] |
| Avidan (a; ETAG), 2008 | 2 | 449 | 1 | 522 | 974 | 2.27 [0.23, 22.03] |
| Avidan (a; BIS), 2008 | 1 | 450 | 5 | 511 | 967 | 0.30 [0.06, 1.49] |
| Errando, 2008 | 18 | 2303 | 4 | 1676 | 4001 | 2.67 [1.14, 6.24] |
| Stait, 2008 | 6 | 112 | 2 | 198 | 318 | 5.26 [1.23, 22.44] |
| Xu, 2009 | 31 | 5991 | 15 | 5064 | 11101 | 1.70 [0.95, 3.04] |
| Avidan (b), 2011 | 12 | 2401 | 15 | 3285 | 5713 | 1.10 [0.51, 2.35] |
| Wang, 2011 | 13 | 1233 | 8 | 1046 | 2300 | 1.37 [0.58, 3.24] |
| Zhang (BIS), 2011 | 3 | 1653 | 1 | 1236 | 2893 | 2.07 [0.28, 15.00] |
| Zhang (SP), 2011 | 8 | 1301 | 7 | 964 | 2280 | 0.85 [0.30, 2.36] |
| Sebel, 2014 | 43 | 11085 | 28 | 8419 | 19575 | 1.16 [0.73, 1.86] |
| Singla, 2017 | 5 | 599 | 2 | 290 | 896 | 1.20 [0.25, 5.87] |
| Yu, 2017 | 7 | 539 | 7 | 691 | 1244 | 1.28 [0.44, 3.71] |
| Hou, 2019 | 9 | 2764 | 14 | 2717 | 5504 | 0.64 [0.28, 1.44] |
| Heggy, 2020 | 0 | 90 | 1 | 109 | 200 | 0.44 [0.03, 7.23] |
| Lennertz, 2022 | 4 | 228 | 2 | 104 | 338 | 0.91 [0.16, 5.18] |
| Celebioglu (Droperidol), 2022 | 3 | 9 | 2 | 16 | 30 | 2.63 [0.38, 18.06] |
| Peto Model ($Q = 27.09$, $df = 42$, $p = 0.57$; $I^2 = 0.0\%$) | | | | | | 1.38 [1.13, 1.68] |
| MH Method ($Q = 21.13$, $df = 42$, $p = 0.85$; $I^2 = 0.0\%$) | | | | | | 1.39 [1.13, 1.71] |
| DL Method ($Q = 23.11$, $df = 42$, $p = 0.99$; $I^2 = 0.0\%$, $\tau^2 = 0.00$) | | | | | | 1.35 [1.15, 1.58] |
| PM Method ($Q = 23.11$, $df = 42$, $p = 0.99$; $I^2 = 0.0\%$, $\tau^2 = 0.00$) | | | | | | 1.35 [1.15, 1.58] |
| REML Method ($Q = 23.11$, $df = 42$, $p = 0.99$; $I^2 = 0.0\%$, $\tau^2 = 0.00$) | | | | | | 1.35 [1.15, 1.58] |
| REML Method, ad hoc SE adj. ($Q = 23.11$, $df = 42$, $p = 0.99$; $I^2 = 0.0\%$, $\tau^2 = 0.00$) | | | | | | 1.35 [1.09, 1.67] |
| REML Method, z-distribution SE ($Q = 23.11$, $df = 42$, $p = 0.99$; $I^2 = 0.0\%$, $\tau^2 = 0.00$) | | | | | | 1.35 [1.10, 1.66] |
| GLMM (CM, AL) ($Q$ (LRT) $= 34.58$, $df = 29$, $p = 0.22$; $I^2 = 0.0\%$, $\tau^2 = 0.00$) | | | | | | 1.38 [1.12, 1.69] |

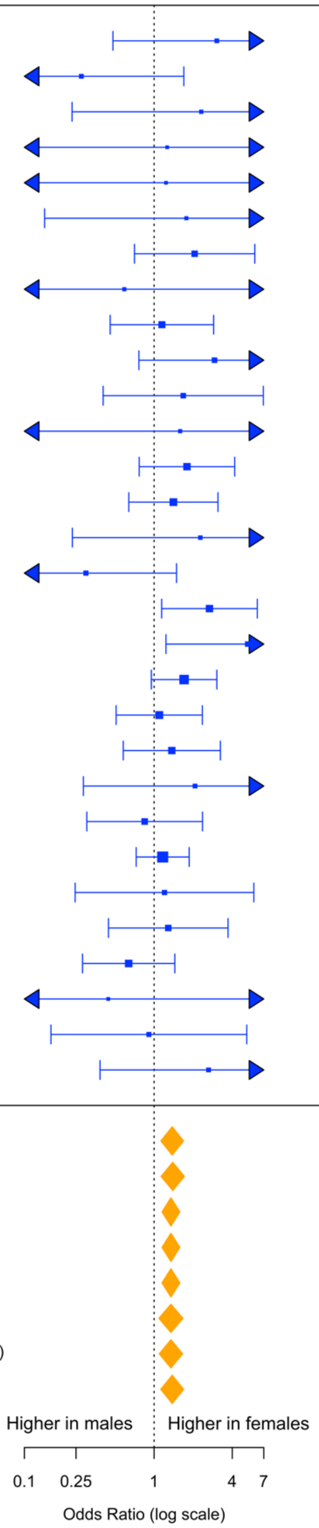


**Supplementary figure 3.** Summary forest plot of total (confirmed and possible) awareness data for differing methods of analysis. BIS, Bispectral index; ETAG, end-tidal anaesthetic gas; 0.2mg/kg, etomidate 0.2mg/kg; 0.3mg/kg, etomidate 0.3mg/kg; 0.4mg/kg, etomidate 0.4mg/kg; M-15, midazolam 15mcg/kg; M-30, midazolam 30mcg/kg; M-45, midazolam 45 mcg/kg; Prop, propofol; PLAC, placebo; RC, routine care; remi, remifentanil; sevo, sevoflurane.

| Authors, Year | Females Aware | Females Unaware | Males Aware | Males Unaware | n | Study Weight | Odds Ratio (95% CI) |
|---|---|---|---|---|---|---|---|
| Miller (Midaz-45), 1996 | 1 | 15 | 0 | 6 | 22 | 0.37% | 1.23 [0.05, 27.90] |
| Miller (Midaz-15), 1996 | 1 | 17 | 0 | 7 | 24 | 0.38% | 1.26 [0.06, 27.83] |
| Lennertz, 2022 | 1 | 231 | 0 | 106 | 338 | 0.41% | 1.35 [0.07, 26.83] |
| Heggy, 2020 | 0 | 90 | 1 | 109 | 200 | 0.47% | 0.44 [0.03, 7.23] |
| Nordstrom, 1997 | 2 | 716 | 0 | 282 | 1000 | 0.76% | 1.77 [0.14, 21.94] |
| Liu, 1999 | 1 | 623 | 1 | 375 | 1000 | 0.89% | 0.59 [0.03, 10.31] |
| Myles (BIS), 2004 | 1 | 472 | 1 | 747 | 1225 | 0.90% | 1.61 [0.09, 27.69] |
| Ekman, 2004 | 1 | 1928 | 1 | 3015 | 4945 | 0.90% | 1.59 [0.09, 27.21] |
| Avidan (a; ETAG), 2008 | 1 | 450 | 1 | 522 | 974 | 0.94% | 1.16 [0.07, 18.72] |
| Avidan (a; BIS), 2008 | 0 | 451 | 2 | 514 | 967 | 0.94% | 0.30 [0.03, 2.90] |
| Miller (PLAC), 1996 | 4 | 16 | 0 | 5 | 21 | 1.06% | 2.31 [0.23, 22.82] |
| Singla, 2017 | 2 | 602 | 1 | 291 | 896 | 1.24% | 0.97 [0.09, 10.84] |
| Wennervirta, 2002 | 4 | 2946 | 0 | 891 | 3841 | 1.35% | 2.10 [0.26, 16.76] |
| Wilson, 1975 | 4 | 314 | 0 | 172 | 490 | 1.71% | 3.04 [0.48, 19.22] |
| Goldman, 1987 | 0 | 7 | 7 | 16 | 30 | 1.88% | 0.27 [0.04, 1.69] |
| Ranta, 1998 | 8 | 1996 | 2 | 606 | 2612 | 3.36% | 1.20 [0.28, 5.22] |
| Stait, 2008 | 6 | 112 | 2 | 198 | 318 | 3.45% | 5.26 [1.23, 22.44] |
| Kerssens, 2003 | 5 | 20 | 4 | 27 | 56 | 3.59% | 1.68 [0.40, 6.95] |
| Myles (SP), 2004 | 5 | 449 | 6 | 778 | 1238 | 4.79% | 1.46 [0.43, 5.02] |
| Yu, 2017 | 7 | 539 | 7 | 691 | 1244 | 6.45% | 1.28 [0.44, 3.71] |
| Sandin, 2000 | 12 | 7041 | 7 | 4725 | 11785 | 8.62% | 1.15 [0.46, 2.87] |
| Errando, 2008 | 18 | 2303 | 4 | 1676 | 4001 | 10.08% | 2.67 [1.14, 6.24] |
| Sebel, 2014 | 16 | 11112 | 9 | 8438 | 19575 | 11.58% | 1.34 [0.61, 2.96] |
| Avidan (b), 2011 | 12 | 2401 | 15 | 3285 | 5713 | 12.39% | 1.10 [0.51, 2.35] |
| Xu, 2009 | 31 | 5991 | 15 | 5064 | 11101 | 21.49% | 1.70 [0.95, 3.04] |
| Peto Model (Q = 17.99, df = 33, p = 0.80; $I^2$ = 0.0%) | | | | | | 100.00% | 1.52 [1.16, 1.99] |

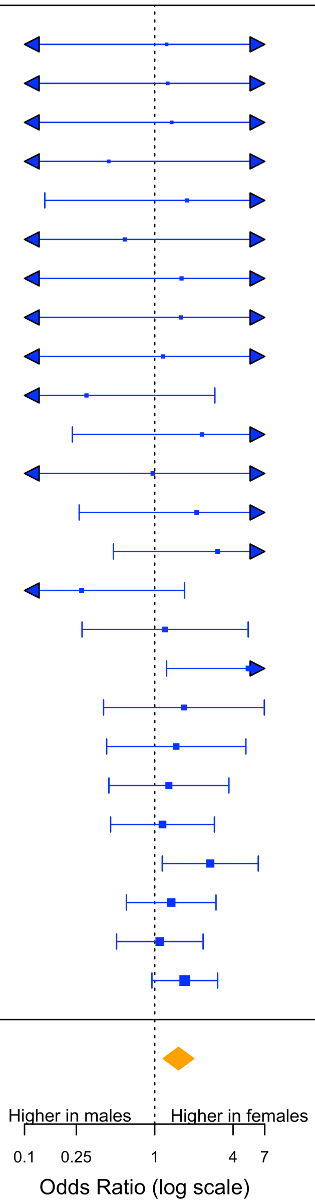


**Supplementary figure 4.** Forest plot of confirmed incidence of awareness with recall with removal of studies of concern from REAPPRAISED assessment. BIS, Bispectral index; ETAG, end-tidal anaesthetic gas; Midaz-15, midazolam 15mcg/kg; Midaz-30, midazolam 30mcg/kg; Midaz-45, midazolam 45 mcg/kg; PLAC, placebo; SP, standard practice.

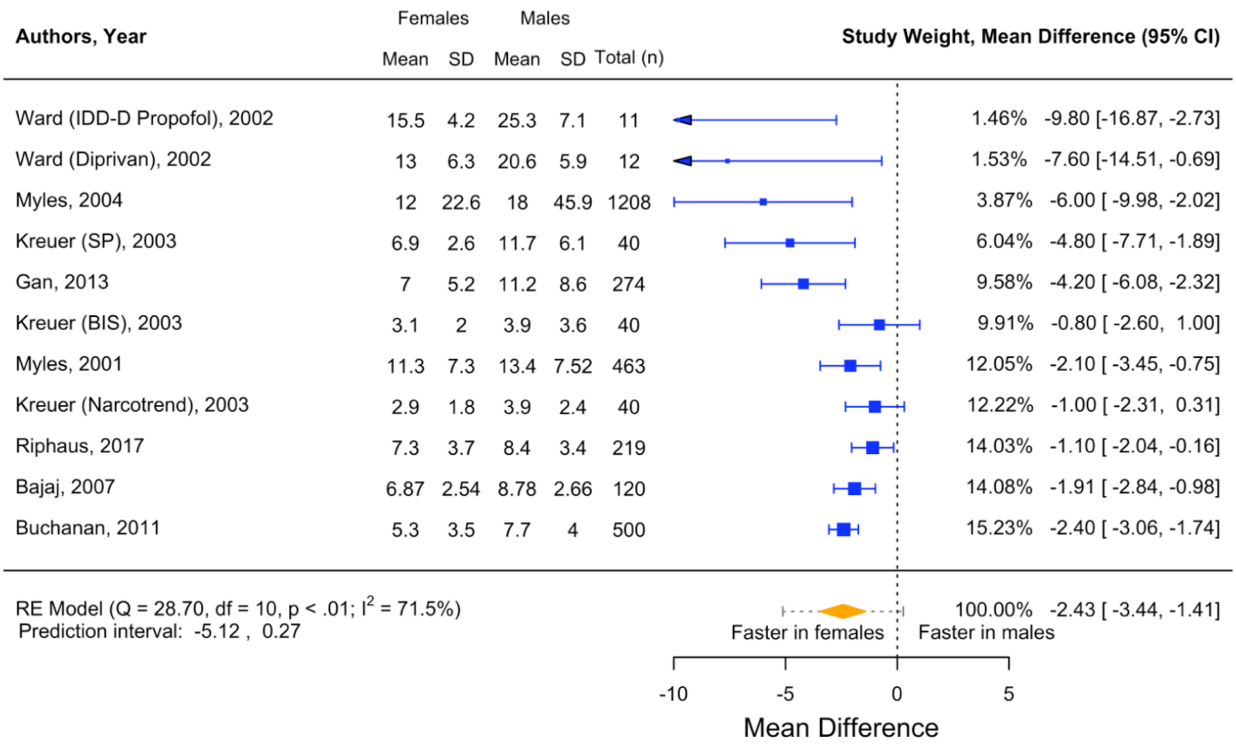


**Supplementary figure 5.** Forest plot of emergence times (time to eye opening) in females versus males with exclusion of studies of concern from REAPPRAISED assessment. BIS, Bispectral Index; SP, standard practice.

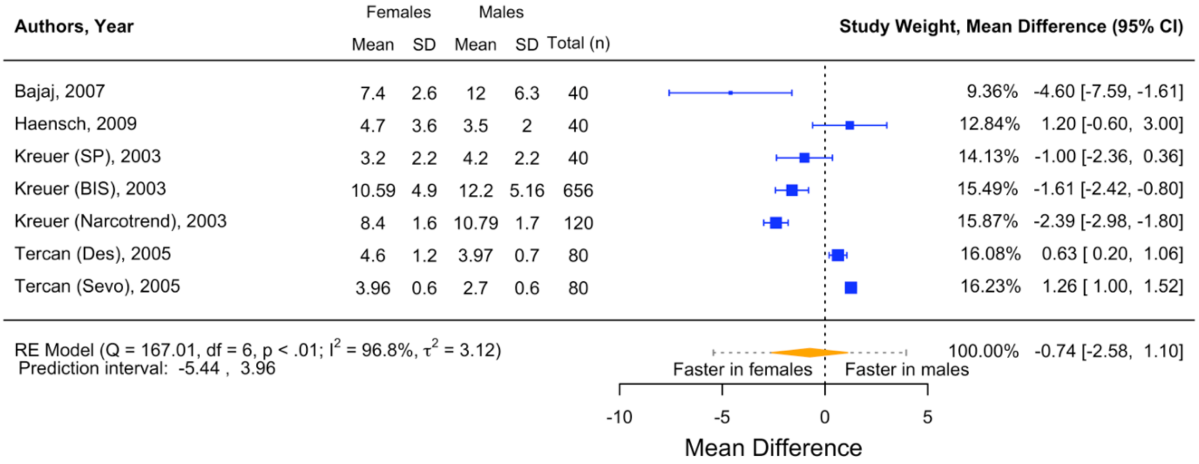


**Supplementary figure 6**. Forest plot of emergence times (time to extubation) in females versus males. BIS, bispectral index; Des, desflurane; Sevo, sevoflurane; SP, standard practice.
